# Shared Risk Genes and Casual Relationships across Sex Hormone Related Traits and Alzheimer’s Disease

**DOI:** 10.64898/2026.04.23.26351626

**Authors:** Chenyu Yang, Noah Cook, Youjie Zeng, Sathesh K. Sivasankaran, FinnGen, Alex DeCasien, Shea J Andrews, Michael E. Belloy

## Abstract

**Background:** Alzheimer’s disease (AD) exhibits marked sex differences. While sex hormone levels across the lifespan likely contribute to this, little remains known about their causal impact and their relation to sex-biased genetic risk for AD. We therefore sought to identify potential shared genetic architectures, as well as causal genes and relationships, between sex hormone-related traits and AD risk.

**Methods:** Large-scale AD sex-stratified genome-wide association study (GWAS) results were available from case-control, proxy-based, and population-based cohorts, including the Alzheimer’s Disease Genetics Consortium, Alzheimer’s Disease Sequencing Project, UK Biobank, and FinnGen. Sex hormone-related trait GWAS were available for age at menarche, menopause, and voice breaking, as well as testosterone, sex hormone-binding globulin (SHBG), progesterone, follicle stimulating hormone, luteinizing hormone, and estradiol levels. Cross-trait conjunctional analyses were conducted to identify pleiotropic overlap between sex-hormone traits and AD, followed by prioritization of candidate causal sex-biased AD genes through quantitative trait locus genetic colocalization analyses. The potential regulatory impact of sex hormones on these genes was assessed through transcription factor motif analyses. Finally, sex-stratified mendelian randomization analyses were used to infer causal effects of sex hormones on AD risk.

**Results:** Genome-wide pleiotropy analyses demonstrated enrichment of AD with testosterone, SHBG, and age-at-menarche traits in women. We identified 12 high-confidence pleiotropic loci, 9 of which showed stronger AD effect sizes in women (3 in men) and 8 that were novel. Genes at these loci were often causally implicated in brain tissues and enriched for promoter-associated androgen receptor transcription factor binding motifs. Mendelian randomization indicated higher bioavailable testosterone in women (OR:0.88; 95%-CI:0.82–0.96) and higher SHBG levels in men (OR:0.86; 95%-CI:0.77–0.96) were associated with lower AD risk.

**Conclusions:** Our findings reveal sex-specific shared genetic architectures between AD and sex hormone-related traits and nominate related genes that may drive sex-biases in AD risk. Several of the implicated female-biased genes are relevant to phosphatidylinositol and lipid metabolism, including *Fatty Acid Desaturase 2* (*FADS2*). While we observed no causal effect of estradiol-related traits on AD risk, the protective effects of bioavailable testosterone in women and SHBG in men provide targets for sex-informed AD risk stratification and prevention strategies.

## Background

Sex differences are pervasive in Alzheimer’s disease (AD) and are evident at the levels of female-biased disease prevalence and incidence, as well as clinical heterogeneity, pathological burden, brain resilience, and response to therapy [1–8] . Growing evidence indicates sex-biases in the genetics of AD contribute to this dimorphism, including the *APOE*\*4 allele, the strongest common genetic risk factor for AD which confers higher risk in women[9–11], as well as several other sex-biased risk genes across the genome [12–15] . Other converging findings indicate sex hormones and chromosomes play a profound role in AD pathobiology and brain resilience[16,17]. While many studies have focused on understanding the roles of menopause and hormone replacement therapy in female-biased AD risk, less is known about how intrinsic estradiol levels or exposure across the lifespan shape risk, and even less about the role of other sex hormones such as testosterone and sex hormone-binding globulin (SHBG) in either female or male-biased disease etiology. Importantly, sex hormones may also interact with genetic risk for AD or even affect expression of risk genes through sex hormone response elements to shape sex-biases in risk. Altogether, it has become evident that a better understanding of the impact of sex hormone biology on AD is crucial to advance sex-informed precision medicine[18].

A core challenge precluding novel insights is the need for structured measurement of sex hormones in large-scale AD-relevant cohorts to allow for causal inference. Some of these hurdles can however be circumvented using a genetics-centered approach to elucidate causal mechanisms. Recent years have seen the advent of large-scale sex-stratified GWAS of sex hormones, as well as age-at-menarche and age-at-menopause as proxies for lifetime estradiol levels or exposure, and confirmed these traits display significant heritability [19–22] . Just recently, a novel sex-stratified AD GWAS was completed in nearly 1 million individuals–representing a substantive expansion in scale compared to prior such efforts–revealing many sex-biased genetic signals and risk genes[15]. Here, we leveraged these novel resources to probe genetic overlap, i.e. pleiotropy, between sex hormones and AD, using a sex-informed framework to pinpoint genetic risk loci that may confer sex-biases to AD risk due to their overlap with sex hormone genetics. We next performed computational genomics analyses at these loci to prioritize likely causal, sex-biased AD risk genes and determine whether their expression may be impacted by sex hormones. Finally, we leveraged Mendelian randomization analyses to investigate whether genetically predicted sex hormone levels causally affect AD risk.

## Methods

Extended methodological details are provided in the **Supplement Methods**. Participants or their caregivers provided written informed consent in the original studies. The study protocol was granted an exemption by the Washington University Institutional Review Board because the analyses were carried out on “de-identified, off-the-shelf” data; therefore, additional informed consent was not required. The FinnGen ethics statement is available in the supplement (Nr HUS/990/2017).

### Genetic and Phenotypic Data Ascertainment

Large-scale AD sex-stratified GWAS meta-analysis results were available from case-control, proxy-based, and population-based cohorts, including the Alzheimer’s Disease Genetics Consortium [23] and Alzheimer’s Disease Sequencing Project [24] (stage-1, clinically diagnosed cases and controls), UK Biobank[25] (UKB, stage-2, family-history proxy diagnoses of AD or dementia), and FinnGen[26] (stage-3, health registry-based AD diagnoses), comprising a total of 636,272 women and 507,902 men. These data are described in detail in Cook *et al. [15]* Sex-stratified sex hormone-related trait GWAS results were available for age-at-menopause [20] , age-at-menarche [21] , age-at-voice breaking [22] , total and bioavailable testosterone levels, and SHBG levels, with and without adjustment for body mass index (BMI) [19] , as well as progesterone, follicle stimulating hormone (FSH), luteinizing hormone (LH), and estradiol levels [27] . The former traits were derived primarily from UKB as well as datasets from the Reproductive Genetics Consortium, Breast Cancer Association Consortium, and Ovarian Cancer Association Consortium, with sample sizes ranging ∼150,000 to ∼550,000[19–22]. The progesterone, FSH, LH, and estradiol GWAS were derived from sex-specific meta-analyses across UKB, ALSPAC, deCODE, EstBB and G&H, with sample sizes ranging ∼6,000 to ∼93,000 individuals [27] . As a negative control for pleiotropy analyses, we leveraged sex-stratified natural hair color GWAS from UKB (manifest release 20180731) [25] . The GWAS datasets leveraged in the present study were derived from individuals of European ancestry.

### Genetic pleiotropy statistical analysis

To assess sex-stratified pleiotropic enrichment across sex hormone-related traits and AD, we used conditional quantile-quantile (QQ) and fold enrichment plots. These visualize genome-wide pleiotropy patterns for AD conditioned on respective sex hormone-related traits at three significance cut-offs (P_Horm_<0.1, P_Horm_<0.01, P_Horm_<0.001). In other words, these plots summarize the distribution of AD association P-values among variants increasingly associated with the hormone-related trait compared to all variants. For these analyses, we used AD GWAS summary statistics excluding UKB to avoid sample overlap with sex hormone-related trait GWAS.

To identify shared, pleiotropic loci between each sex hormone-related trait and AD, we conducted pairwise conjunctional false discovery rate (FDR) analyses following a previously established framework[28,29]. Briefly, conjunctional FDR (conjFDR) is an extension of conditional FDR that seeks to assess whether a genetic variant is associated with one trait given its association with another–formally, for a given variant, it is the posterior probability that the variant is null for the primary trait, conditional on its observed P-values in both traits, with ConjFDR computed as the maximum of the two directional conditional FDRs (primary trait conditioned on the secondary trait and vice versa). Variants with conjFDR<0.05 were determined to be pleiotropic [28] . For these analyses, we used AD GWAS results including all cohorts to increase discovery power. To verify robustness of resultant loci, we confirmed consistency of AD effect estimates with and without inclusion of UKB.

### Tier-based prioritization of pleiotropic loci

Pleiotropic variants were grouped into variant sets representing independent signals using linkage disequilibrium (LD; R2<0.01) clumping and annotated with Tier-3 status. We then evaluated which signals contained variants that displayed sex-biased associations with AD risk, requiring P<0.05 on sex-heterogeneity tests (Z - value = (Beta_Men_ - Beta_Women_)/ √ (SE_Men_^2^ + SE_Women_^2^)) or >1.5-fold effect size differences across sexes, to advance them to Tier-2 status. Finally, to identify which signals had evidence of sharing a causal variant between AD and the respective sex hormone-related trait (rather than pleiotropic overlap driven by LD), we performed genetic colocalization analyses (COLOC; cf. supplementary methods) [30,31] . Signals that showed a large posterior probability of sharing a causal variant (PP4≥0.7) were advanced to Tier-1 status. Novelty of Tier-1 loci was annotated considering proximity (<1Mb) to genome-wide significant risk loci in prior large-scale AD GWAS.

### Gene prioritization

To prioritize putative causal genes and epigenetic regulatory features at Tier-1 loci, we performed genetic colocalization analyses (COLOC; cf. supplementary methods)[30,31] between the local AD GWAS signals and a series of quantitative trait locus (QTL) datasets. These included bulk tissue and cell-type specific resources that have mapped variants to gene expression (eQTL), transcript splicing (sQTL), protein abundance (pQTL), methylation (mQTL), chromatin accessibility (caQTL), and histone acetylation (haQTL) features (**Table-S1**). Strong colocalization was defined as PP4≥0.70 and used to nominate candidate genes and features.

### Sex hormone transcription factor analyses

For genes prioritized at Tier-1 loci, we performed *de novo* transcription factor motif enrichment analysis on promoter sequences using HOMER[32]. Promoter regions were defined as -1000 to +300 bp around the annotated transcription start site of each gene. Female-biased and male-biased genes were analyzed separately. Identified *de novo* motifs were annotated by similarity to known transcription factor binding motifs from the HOCOMOCO v11 collection [33] . These analyses were conducted to assess whether candidate sex-biased genes showed enrichment of promoter-associated androgen or estrogen receptor-related motifs. Motifs were filtered to those having at least P-values<0.05.

Additionally, we used motifbreakR to evaluate whether variants at Tier-1 loci directly overlapped predicted androgen receptor (AR) or estrogen receptor (ESR1/ESR2) transcription factor binding motifs and altered motif match scores between alleles[34,35]. Analyses were performed using curated AR, ESR1, and ESR2 position weight matrices from JASPAR, HOCOMOCO, and SwissRegulon [34,35] . Following default guidelines, motif matches were scaled from 0 to 1 as a fraction of the possible score range for each motif, and variants were retained when either the reference or alternate allele exceeded a score threshold of 0.85.

### Two-sample Mendelian randomization statistical analysis

To investigate potential causal effects of sex hormone-related traits on AD risk, we conducted sex-matched two-sample Mendelian randomization (MR) analyses using sex hormone-related traits as exposures and AD as the outcome. To increase power, the AD GWAS in these analyses included all cohorts. For each exposure (sex hormone-related trait), we selected independent (R2<0.001) strongly associated (P<5e-8) variants as instrumental variables. Variant-exposure and variant-outcome association estimates were aligned to the same effect allele and ambiguous variants were excluded. The primary MR estimator was the inverse-variance-weighted (IVW) method under a random-effects model. Per sex, we then identified sex hormone-related traits significantly associated with AD (FDR-P<0.05). For significant associations, we carried out a series of sensitivity checks. Consistency of effect sizes and association significances was evaluated with three additional MR approaches: MR-Egger, weighted median, and weighted mode, while horizontal pleiotropy was evaluated using the MR-Egger intercept and MR-PRESSO global test[36–39]. Variant heterogeneity and the influence of individual variants were evaluated using Cochran Q statistics under IVW and MR-Egger models and leave-one-out analyses. We additionally re-estimated IVW MR effects using procedures that adjust standard errors for sample overlap between exposure and outcome GWAS due to shared UKB participants. To examine whether the male-stratified association between SHBG and AD was independent of BMI, we performed multivariable MR including SHBG and BMI as joint exposures and AD as the outcome. All MR analyses were implemented in R using standard packages[40–42].

## Results

An overview of the study design is shown in **Figure-1**. In assessing pleiotropy at a genome-wide level, women displayed enrichment of AD associations signals when variants were conditioned on their associations with bioavailable and total testosterone, SHBG, and age-at-menarche, but not age-at-menopause, estradiol, progesterone, follicle stimulating hormone, or luteinizing hormone (**Figure-2**; **Figure-S1**). In contrast, men showed weaker or no enrichment for any of the sex hormone-related traits. Conditioning on hair color as a negative control showed no evidence of AD enrichment in either sex (**Figure-S2**), supporting the specificity of our findings.

**Figure 1.**
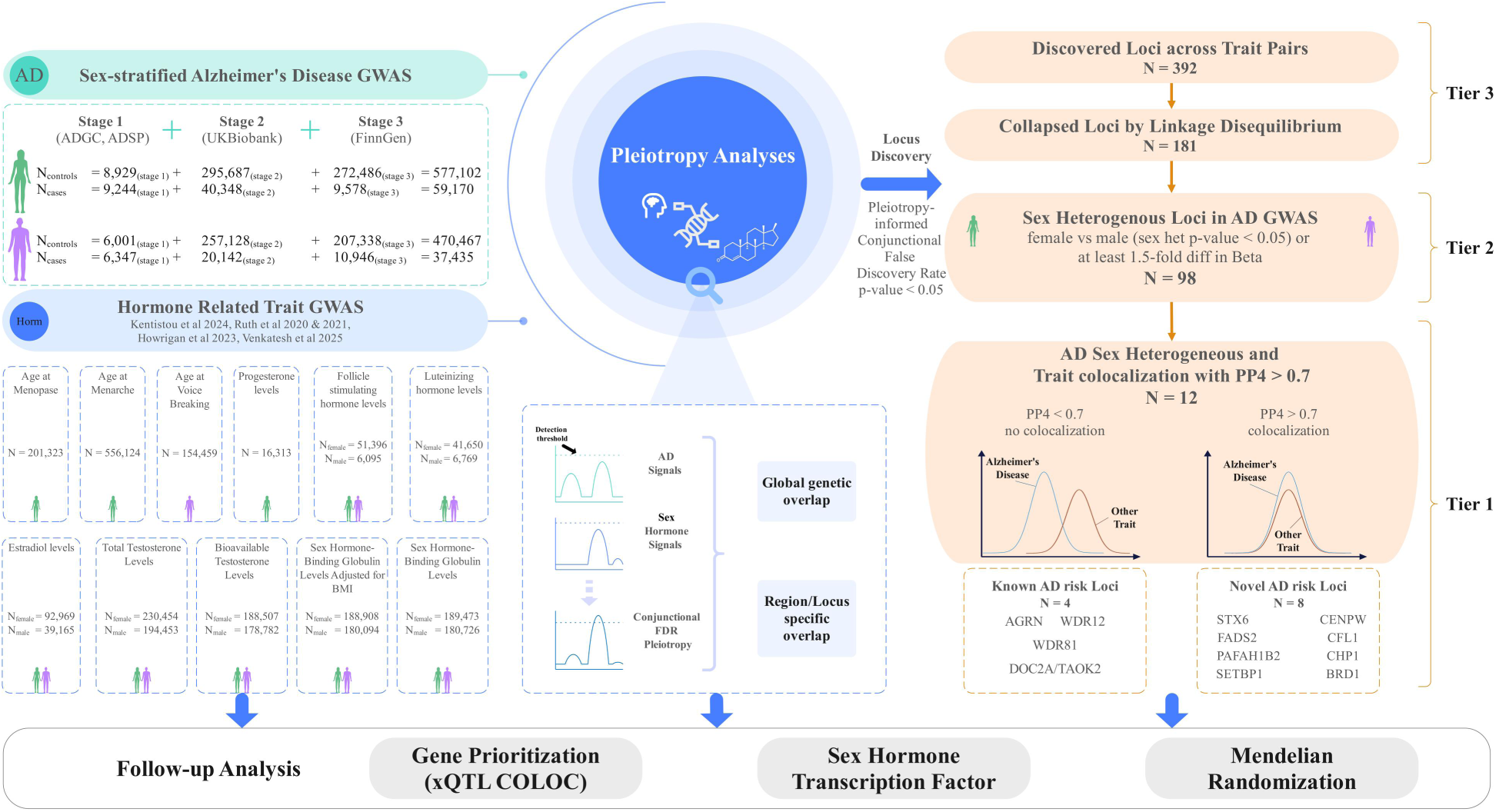
Schematic overview of the study. **Left:** Sex-matched pleiotropic and causal relationships across 7 sex-hormone related trait and Alzheimer’s disease (AD) genome-wide association studies (GWAS) were evaluated. **Center:** Genetic pleiotropy was quantified through the conjunctional false discovery rate (conjFDR) approach, assessing (i) global genome-wide enrichment and (ii) regionally shared signals. **Right:** Pleiotropic loci were filtered following a tiered prioritization approach: Tier 3, locus displays significant pleiotropy (conjFDR<0.05); Tier 2, locus additionally displays sex heterogeneity in the AD GWAS; Tier 1, locus further shows evidence of sharing an underlying causal variant across AD and the sex hormone related trait (genetic colocalization PP4>0.7). **Bottom:** Follow-up analysis of 12 Tier 1 loci aimed to identify genes that may causally contribute to sex differences in AD (xQTL colocalization) and whether these genes display evidence of sex hormone response element enrichment. Finally, genetic Mendelian randomization analyses were used to probe putative causal effects of sex hormones onto AD.

**Figure 2.**
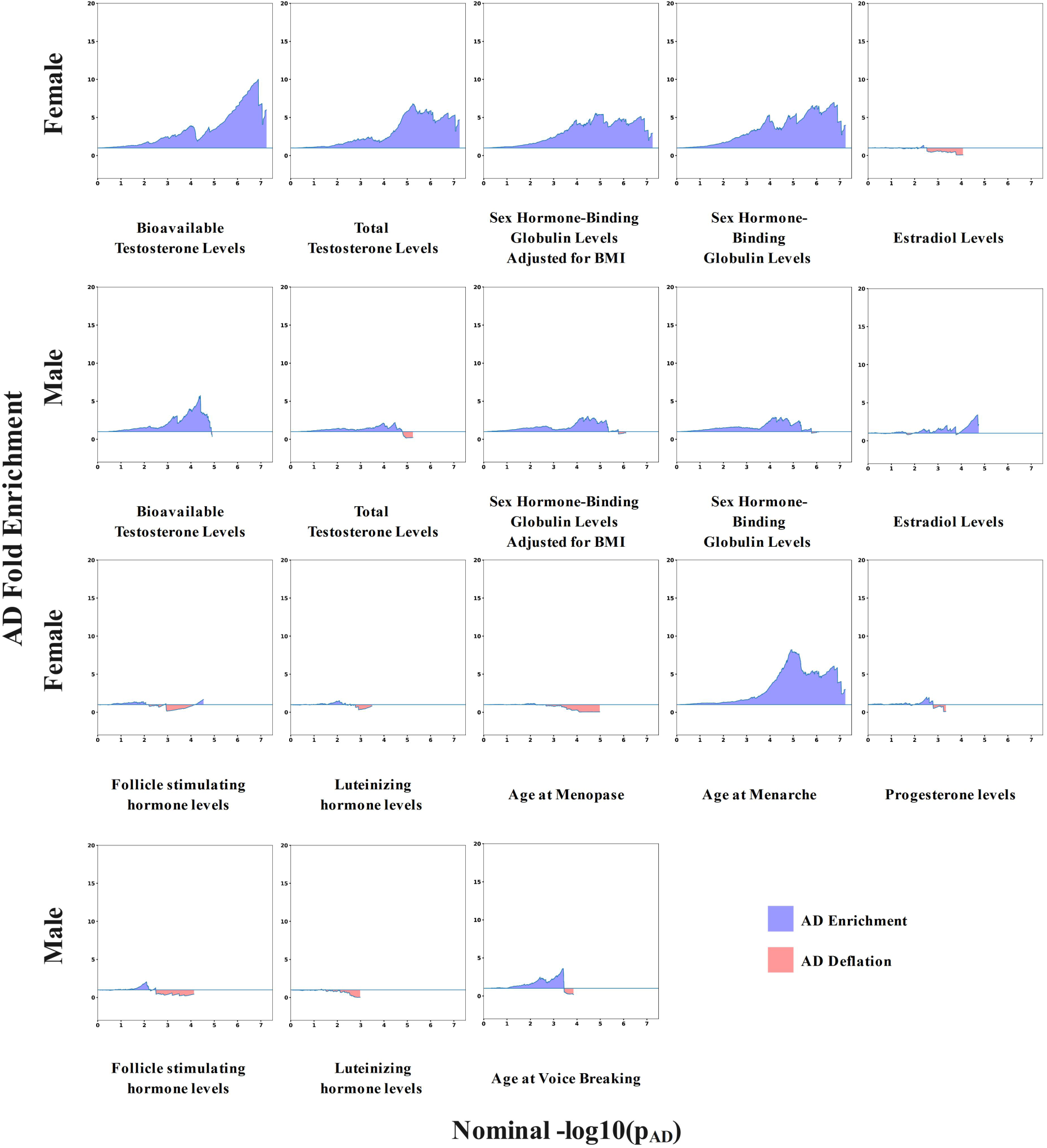
Genome-wide pleiotropy across sex-hormone related traits and Alzheimer’s disease. Plots show enrichment results of Alzheimer’s disease (AD) genetic signals conditioned on sex-hormone related traits in female-matched (rows 1 and 3) and male-matched (rows 2 and 4) analyses. The x-axes indicate significance quantiles of the AD GWAS association findings (-log_10_(P)) while the y-axes indicate fold-enrichment of variants displaying at least suggestive associations (P<1e-3) in the respective sex-hormone related traits relative to all available variants (i.e. the reference; fold enrichment = 1). Positive AD enrichment is marked by blue shaded areas, reflecting pleiotropy, while AD deflation or lack of pleiotropy is marked by red shaded areas. All plots are shown with consistent axes limits, indicating that multiple sex-hormone related traits, except age at menopause, showed larger pleiotropy with AD in women relative to men.

When assessing variant-specific pleiotropy across all trait pairs, conjFDR analyses identified 392 pleiotropic associations corresponding to 181 independent variant sets, i.e. signals (**Table-S2**; circos plots summarizing all pleiotropic discoveries are in **Figure-S3)**. Among these Tier-3 signals, 98 contained variants that showed evidence of sex-biased effects in AD GWAS (Tier-2), while 12 of those 98 signals further showed evidence of cross-trait genetic colocalization (Tier-1, **Table-S3**). Of the 12 Tier-1 loci, 8 were novel compared to prior AD GWAS while 9 contained variants that displayed female-biased AD risk associations (**Figure-1; Figure-3A**). AD effect estimates at all 12 Tier-1 loci were highly consistent with and without inclusion of UKB samples, supporting AD signal robustness (**Figure-S4**). As an illustration of the types of discoveries enabled by our approach, we showcase the novel female-biased *FADS2* locus in **Figure-3B-C**. Detailed locus zoom plots of all Tier-1 loci are provided in **Figure-S5**. It should be noted that since the Tier-1 discoveries represented sets of pleiotropic variants, it was possible that the best colocalizing variant at a respective locus–i.e. the most likely causally shared variant across the trait pair–did not display evidence of sex-biased effects in AD GWAS contrary to other variants in the set. This was the case for 3 out of 12 Tier-1 loci, *DOC2A/TAOK2*, *WDR81*, and *STX6*, indicating these are less robust in terms of their potential impact on sex-biased AD risk compared to the other 9 Tier-1 loci.

**Figure 3.**
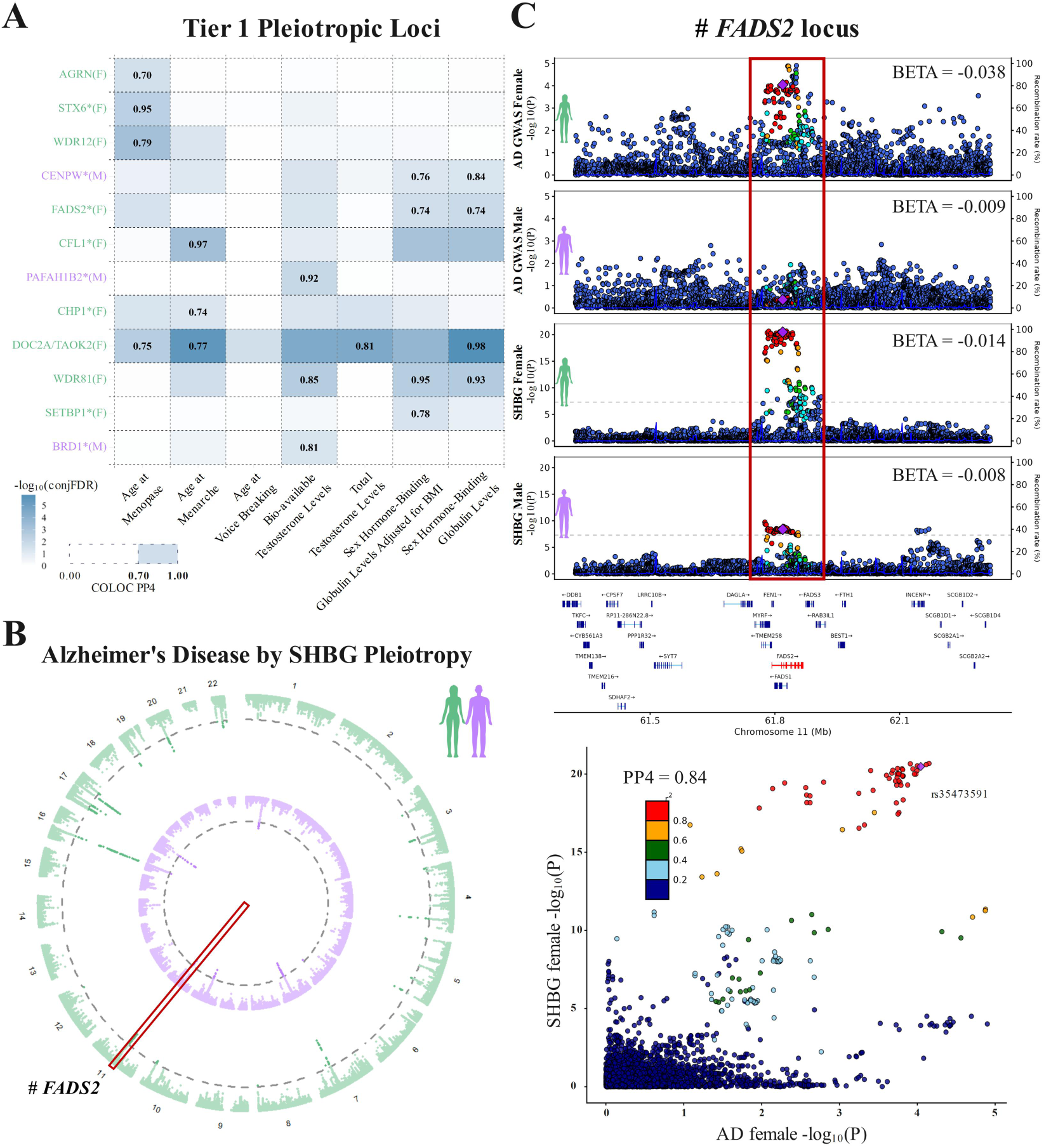
Overview of prioritized pleiotropic loci between Alzheimer’s disease and sex-hormone related traits. **A)** Matrix of Tier 1 pleiotropic loci. Rows represent independent signals, annotated with the nearest gene or known Alzheimer’s disease (AD) locus and whether it showed a female (F) or male (M) biased association with AD. Columns represent which sex-hormone related traits were compared with AD. Cell colors indicate the pleiotropy significance at the locus (-log_10_(conjFDR)). Results from genetic colocalization analyses (COLOC) are shown only for loci and trait pairs with conjFDR<0.05 and COLOC PP4>0.7, reflecting strong support for a shared causal variant. **B)** Circos plot illustrating pleiotropic loci across the sex-hormone binding globulin (SHBG) levels GWAS and Alzheimer’s disease GWAS in female-matched (outer ring, green) and male-matched (inner ring, purple) analyses. The dotted circular lines mark conjFDR<0.05. The red box denotes the **C)** *FADS2* Tier 1 locus. Top to bottom: Locus zoom plots for female AD GWAS, male AD GWAS, female SHBG GWAS, and male SHBG GWAS. Y-axes show -log_10_(P) in the corresponding GWAS. Beta coefficients across panels indicate the tagging (diamond) variant associations in respective groups and traits. Dot colors mark LD with the tagging variant (cf. legend). Bottom: Locus-compare plot showing the AD female (x-axis) and female SHBG signals (y-axis) colocalize (COLOC PP4=0.74).

The identified Tier-1 signals represented the top candidate loci that may contribute to sex differences in genetic risk for AD due to their genetic overlap with sex hormone-related traits. We therefore leveraged QTL colocalization analyses to identify which potential causal genes and epigenetic features they regulate. Detailed findings are provided in **Figure-S6** and **Table-S4**, while top gene prioritizations are shown in **Figure-4A**. Regulatory effects were successfully nominated at 11/12 loci. Notably, at 9 loci, genes were implicated in brain bulk tissues or cell-types (in addition to other potentially relevant tissues), showcasing how these loci may contribute to AD risk. Similarly, across 10 loci, 8 displayed evidence of regulating methylation in brain while 5 related to microglial chromatin accessibility (**Figure-S6**). To understand how the Tier-1 loci could contribute to sex biases in AD risk, we investigated if their prioritized genes showed evidence of being regulated by sex hormones (**Figure-S7**). Transcription factor motif enrichment for female-biased genes revealed a significant over-representation of an androgen receptor-like binding motif (P=1e-9; **Figure-4B**), suggesting that at least a subset of loci could be sex-biased due to testosterone biology. Additionally, we found direct evidence that AD-associated variants in the *DOC2A/TAOK2* and *WDR81* locus fell within known androgen and estrogen receptor motifs (**Figure-S8, Table-S8**). Notably, despite that the top colocalizing variants at these two loci did not appear to have sex-biased effects in AD GWAS (cf. above), those that fell within the motif sites did, reaffirming the potential impact of these loci on sex-biased AD risk.

**Figure 4.**
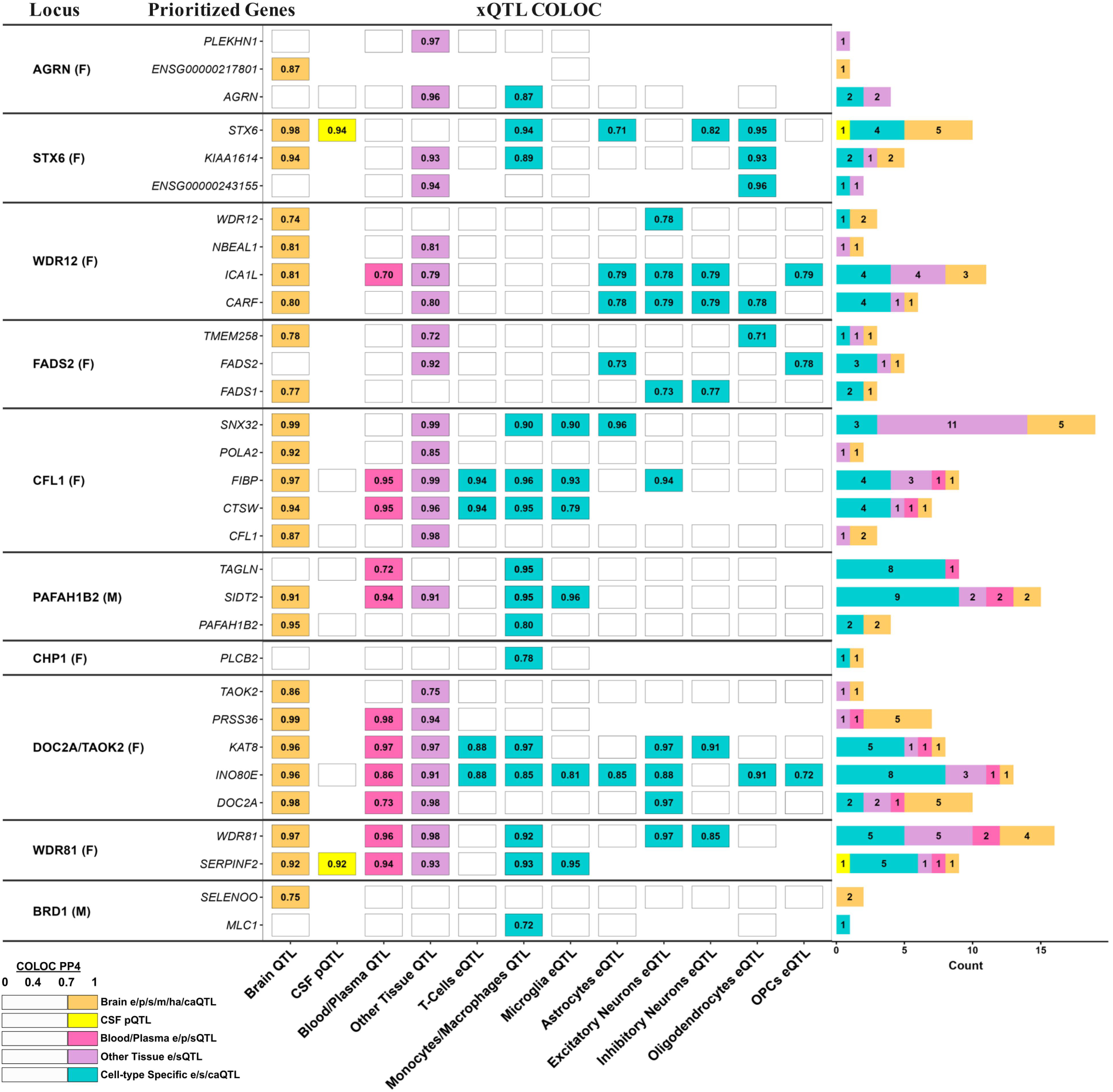
Overview of gene prioritization and motif enrichment at Tier 1 pleiotropic loci. Summary of genetic colocalization (COLOC) analyses to prioritize potential causal genes at Tier-1 pleiotropic loci. Columns reporting findings for different QTL types (xQTL) across different tissues and cell-types. Top supported genes are listed per locus. For each category, the cell reports the best PP4 among the datasets collapsed within that category (e.g., “Other Tissue” corresponds to the best result across GTEx v8 tissues). Only COLOC findings with PP4>0.7 are displayed and contributed to gene prioritization. *Abbreviations: eQTL, expression QTL; sQTL, splicing QTL; pQTL, protein QTL*.

Finally, we sought to investigate if the sex hormone-related traits displayed evidence of sex-stratified causal effects on AD risk (**Figure-5A**). Genetic MR analyses showed that higher bioavailable testosterone was associated with reduced AD risk in women (OR, 0.88; 95% CI, 0.82-0.96; FDR-P=0.013), while higher BMI-adjusted SHBG was associated with reduced AD risk in men (OR, 0.86; 95% CI, 0.77-0.96; FDR-P=0.030) (**Figure-5B-C**). MR sensitivity analyses confirmed consistent effect estimates under different MR models and displayed no evidence of substantial heterogeneity or horizontal pleiotropy (**Figure-S9**, **Table-S5**). MR analyses that accounted for sample overlap with UKB also indicated consistent associations (**Table-S6**). Finally, since the SHBG associated with reduced risk in men was only significant with the BMI-adjusted SHBG GWAS data, we performed multivariable MR analyses including both SHBG and BMI as exposures in men. While SHBG continued to display a protective effect size, it was no longer significant for all MR estimators, indicating the SHBG finding may be less robust (**Table-S7**).

**Figure 5.**
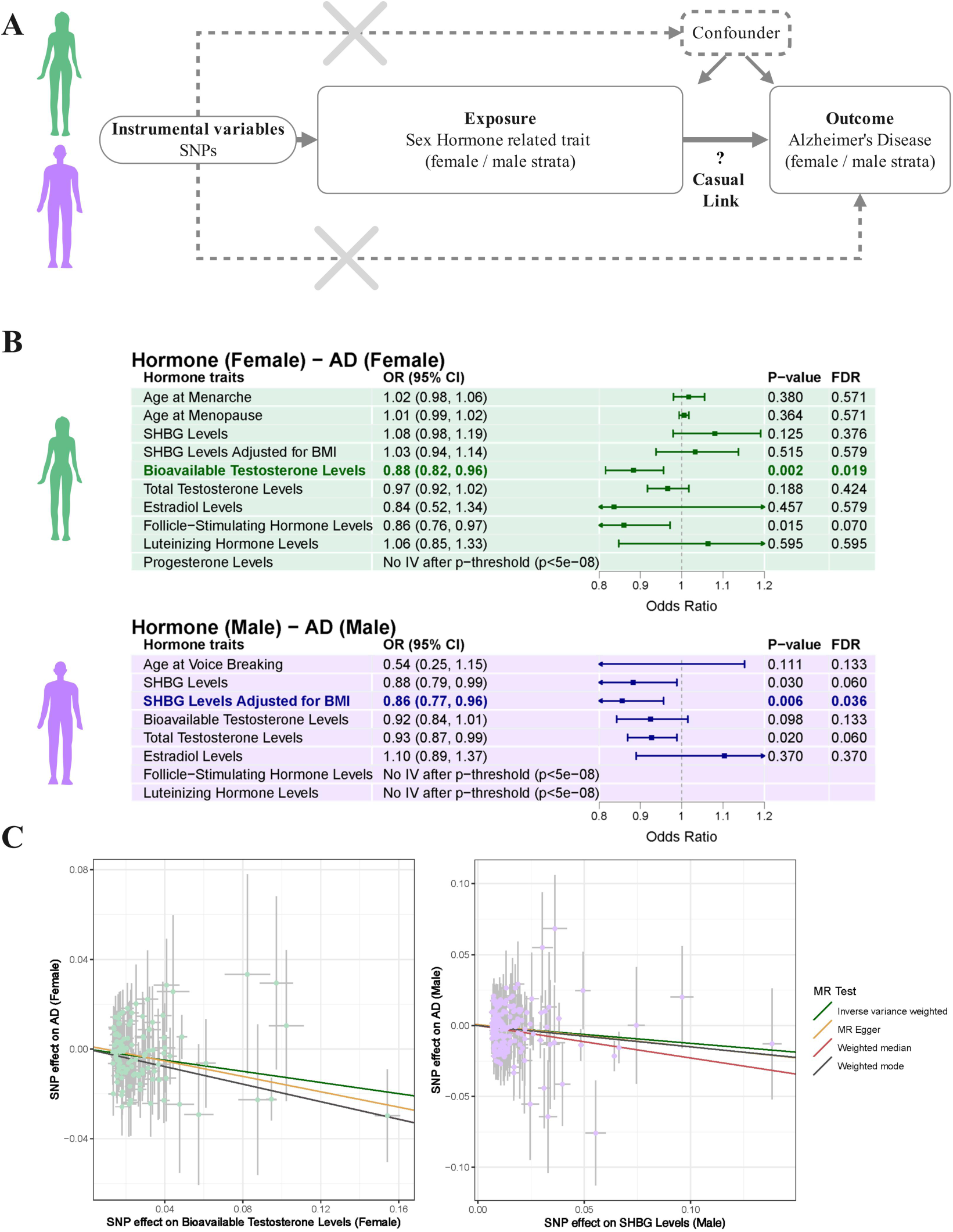
Causal effects of x hormone-related traits on Alzheimer’s disease risk evaluated by Mendelian randomization. **A)** Schematic of the sex-matched two-sample Mendelian randomization (MR) analysis design. **B)** Forest plots show sex-matched MR findings. Points denote odds ratios (OR) with 95% CIs from the inverse-variance weighted (IVW) MR test. Reported alongside each trait are the P-value and FDR-corrected P-value. Two trait-AD pairs with FDR-significant associations are highlighted while matching **C)** MR scatter plots show variant effects on the exposure (sex-hormone related trait, x-axis) versus variants effects on the outcome (AD, y-axis). Variant effects are reported as beta coefficients while error bars provide standard errors. Colored lines indicate the slopes from different MR tests, including the primary IVW analysis, as well as three sensitivity analyses (MR-Egger, weighted median, weighted mode). Negative slopes indicate higher sex hormone levels reflect a protective effect on AD risk.

## Discussion

Our study represents the largest-to-date genetic investigation of pleiotropic and causal relationships between sex hormone biology and AD risk. Genome-wide pleiotropy findings indicated that bioavailable and total testosterone, SHBG, and age-at-menarche displayed larger enrichment of AD signals in women than in men. Concordantly, 9/12 prioritized pleiotropic loci displayed female-biased associations with AD. Despite that sex hormone-related trait GWAS pertained to peripheral sex hormone levels, subsequent gene prioritization at pleiotropic loci implicated potential causal genes in brain tissues and cell-types. These findings are concordant with converging evidence that sex-biased AD risk genes are more often observed in women and brain tissues [43,44] . Many prioritized genes were also implicated in microglia and astrocytes, which have been reported to impact sex-biased inflammatory mechanisms and female-biased effects on AD pathogenesis, potentially driven by estrogen biology [45,46] . While genome-wide pleiotropy findings showed no enrichment for age-at-menopause, 4 female-biased loci were causally shared between AD and age-at-menopause. This discrepancy reflects that genome-wide pleiotropy is agnostic to whether genetic signals are causally shared or overlap due to LD-sharing. Out of 9 female-biased pleiotropic loci, 6 were causally shared with either age-at-menopause or age-at-menarche, while 4 were shared with testosterone or SHBG levels (the *DOC2/TAOK2* locus was shared across both sets of sex hormone traits) suggesting that sex differences in both estrogen and testosterone biology may drive female-biased AD risk genes.

The prioritized pleiotropic loci are anticipated to regulate pleiotropic genes involved in both sex hormone and AD pathogenic pathways. Two mechanisms through which these genes could confer sex biases in AD risk are: (i) their loci are enriched for sex hormone response elements which can skew gene abundance in a sex-biased manner, or (ii) their gene-hormone network interactions affect gene abundance and dynamics in a sex-biased manner. In line with the first point, we observed enrichment of an androgen receptor-like element for female risk genes, further corroborating that testosterone biology may play an important role in female-biased AD risk. The *DOC2A/TAOK2* and *WDR81* loci additionally showed AD signals falling within androgen and estrogen receptor motifs, further corroborating that sex hormones could drive sex-biases in their associations with AD. To identify support for the second point, we reviewed the biological mechanisms of our prioritized genes, focusing on genes with the most abundant colocalization support at each locus considering those are most likely to be causal. Notably, multiple female-biased genes, despite having various biological functions, related to phosphatidylinositol and phosphoinositide 3-kinase (PI3K) metabolism, including *FADS2*, *SNX32*, *PLCB2*, *DOC2A*, and *WDR81*. Phosphatidylinositol metabolic pathways have been implicated in AD pathogenesis as well as estrogen biology, where PI3K interacts with Estrogen Receptor Alpha, highlighting how these genes may display female-biased AD risk associations[47–51]. Notably, *FADS2*, which also plays a broader role in fatty acid metabolism and was novel compared to prior AD GWAS loci, has recently been shown to impact cognitive function, associate with biomarker-positive AD risk, be a key enzyme related to lower unsaturated lipid levels in women, and affect steroidogenesis in the adrenal gland [52–55] . Human proteogenomics and experimental studies, including loss-of-function work in *Drosophila*, have additionally indicated a causal role of *SNX32* in neurite outgrowth and neurodegeneration [56–58] . *DOC2A*, which also affects neurotransmitter release, was only the second most prioritized gene at its locus but was previously prioritized in AD GWAS efforts [59] and has additional genomics support for a role across multiple neurodegenerative diseases including AD [60–62] . *WDR81*, already prioritized in prior AD GWAS[59], is additionally relevant to autophagy and endo-lysosomal trafficking, which are both AD-relevant pathways. For other prioritized genes, their role in sex hormone biology was less evident at the current time, but they appeared relevant to AD. *SITD2* and *STX6* play roles in autophagy, endo-lysosomal trafficking, and intracellular protein trafficking, with *STX6* implicated in Tau pathology across multiple neurodegenerative diseases[63,64]. *AGRN* is a heparan sulfate proteoglycan that was prioritized in prior AD GWAS and implicated in amyloid plaque formation as well as other AD-relevant pathways [65,66] . Finally, *SELENOO* is a selenoprotein, implicating it in selenium metabolism which in turn has been linked to Tau pathology[67].

In addition to mapping pleiotropic genes that may confer sex biases in AD risk, we conducted MR analyses in which the goal was to use genetic proxies to determine whether sex hormone levels across the lifespan causally contribute to AD risk in a sex-biased fashion. A surprising null-finding was that age-at-menopause, age-at-menarche, and estradiol displayed no effect on AD risk in women. This is however consistent with recent work by Oppenheimer *et al*. that performed extensive MR analyses of both estradiol levels and estradiol-related traits with a prior, much smaller sex-stratified AD GWAS[68]. Taking together, the results, in a strict sense, indicate genetic regulation of these traits does not affect AD risk, and thus, more generally, point toward there being no causal effect of lifetime estradiol levels or exposure on AD risk. While the GWAS MR approach has limitations and represents a tool to strengthen causal inference rather than providing definitive proof of causality, these findings do raise the question whether prior epidemiological studies may have faced limitations from confounding factors or reverse causation. As argued by Oppenheimer *et al*., causal effects on AD risk may instead be anticipated for dynamic changes and age-specific effects of estradiol. Menopause may also directly impact AD risk independent of the age-at-onset. Our findings suggest that these questions should be focus points on future studies. In contrast to estradiol-related traits, we observed that bioavailable testosterone levels were associated with a 12% reduction in female AD risk–robust under all sensitivity analyses–and showed a trend for 8% reduction in male AD risk. Several prior studies in men have similarly reported lower testosterone levels are associated with higher risk of dementia[69–71]. Much less is known about testosterone in women, but initial work suggests lower levels could relate to poorer cognitive function in *APOE*\*4 carriers [72,73] . While future studies into the role of testosterone in female AD etiology are warranted, our findings suggest testosterone therapies appear as a promising target for both women and men, while measurement of testosterone levels has potential to aid clinical trial stratification and recruitment. Finally, we observed SHBG levels adjusted for BMI were associated with a 14% reduction in male AD risk, while no similar effect was observed in women. Although this finding did not pass sensitivity analyses in which BMI was adjusted through multivariate MR, a recent study by *Buckley et al.* in independent samples consistently observed higher SHBG levels in healthy elder men were associated with lower Tau pathology in the entorhinal cortex, after adjustment for BMI. As reviewed by *Buckley et al.*, the role of SHBG in AD remains uncertain and there have been variable reports regarding its association with AD risk, which likely relates to age-dependent effects of SHBG levels–across the lifespan SHBG increases in men while displaying a U-curve in women [74–76] . Since SHBG strongly binds testosterone to regulate its availability, the non-significant associations of (bioavailable) testosterone with AD risk or Tau pathology in men suggests an androgen-independent role of SHBG. Some potential mechanisms linking SHBG to AD risk include its role in insulin, lipid, and cardiovascular mechanisms[77]. Notably, modifiable lifestyle factors such as smoking, alcohol, and physical activity can influence SHBG levels[75,77]. In summary, future studies are needed to elucidate how exactly SHBG affects AD risk, but our findings point towards a therapeutic potential and the importance of intervention strategies centered on SHBG to reduce AD risk in men.

## Limitations

While we included sex-stratified GWAS with large underlying sample sizes, these pertained only to European ancestry individuals, limiting generalizability of our findings to other populations. Further, the AD GWAS included biobanks which can reduce specificity to AD pathology [15] . Indeed, some of the genes we prioritized were implicated in additional neurodegenerative disorders beyond AD. It should also be considered that sex-biased effect estimates on AD may partially represent sex-specific differences in disease prevalence or underlying environmental factors rather than biological heterogeneity. Experimental studies are therefore warranted to confirm the exact relevance of prioritized loci and genes to AD pathogenesis and the precise mechanisms through which they relate to sex hormone biology and sex differences in AD. Further, the available GWAS of sex hormone-related traits pertained to cross-sectional measurements or proxies of lifetime hormone exposure, providing only a static snapshot of the changing hormone environment over the lifespan. Studies evaluating temporal changes, particularly at older age, and longitudinal measurements of sex hormones to infer causal mechanisms will be important to corroborate and complement our observations.

## Conclusions

Our findings reveal a sex-specific shared genetic architecture between AD and sex hormone-related traits and nominate related genes that may drive sex-biases in AD risk. Several of the implicated female-biased genes are relevant to phosphatidylinositol and lipid metabolism, including *Fatty Acid Desaturase 2* (*FADS2*). While we observed no causal effect of estradiol-related traits on AD risk, the protective effects of bioavailable testosterone in women and SHBG in men provide targets for sex-informed AD risk stratification and prevention strategies.

## List of abbreviations

genome-wide association study: (GWAS)
sex hormone-binding globulin: (SHBG)
odds ratio: (OR)
Fatty Acid Desaturase 2: (FADS2)
Alzheimer’s disease: (AD)
body mass index: (BMI)
follicle stimulating hormone: (FSH)
luteinizing hormone: (LH)
quantile-quantile: (QQ)
false discovery rate: (FDR)
conjunctional false discovery rate: (conjFDR)
linkage disequilibrium: (LD)
quantitative trait locus: (QTL)
expression quantitative trait locus: (eQTL)
transcript splicing quantitative trait locus: (sQTL)
protein abundance quantitative trait locus: (pQTL)
methylation quantitative trait locus: (mQTL)
chromatin accessibility quantitative trait locus: (caQTL)
Mendelian randomization: (MR)
androgen receptor: (AR)
estrogen receptor: (ESR)
phosphoinositide 3-kinase: (PI3K)

## Data availability

AD GWAS data will be available at: https://www.medrxiv.org/content/10.1101/2025.10.31.25339089v1. Data used in the AD GWAS analyses are available upon application to:

- dbGaP (https://www.ncbi.nlm.nih.gov/gap/)
- NIAGADS (https://www.niagads.org/)
- LONI (https://ida.loni.usc.edu/)
- AMP-AD knowledge portal / Synapse (https://www.synapse.org/)
- Rush (https://www.radc.rush.edu/)
- NACC (https://naccdata.org/)
- UKB (https://www.ukbiobank.ac.uk/)
- FinnGen (https://www.finngen.fi/en)

Sex hormone-related GWAS data are available at:

- Age at Menarche:

https://www.repository.cam.ac.uk/items/8c5f7afb-5fa2-45ea-b52d-4e643bc2a5b7

- Age at Menopause:

https://ega-archive.org/studies/EGAS00001004947

- Age at voice breaking: https://docs.google.com/spreadsheets/d/1kvPoupSzsSFBNSztMzl04xMoSC3Kcx3CrjVf4yBmESU/

edit?gid=227859291#gid=227859291

- Testosterone & SHBG levels: https://www.ebi.ac.uk/gwas/studies/

A table overview of all QTL resources and their public identifiers are indicated in **Table-S1** of the supplement.

## Code availability

The code used to support the findings of this study is publicly available at GitHub: https://github.com/Belloy-Lab/Pleiotropy_analysis_on_AD_and_Sex_Hormone

## Author contributions

C.Y. performed data acquisition, designed analyses, performed analyses, and wrote paper. N.C., Y.Z., S.S. performed analyses. A.D., S.J.A. designed study, supervised analyses. M.E.B. performed data acquisition and analyses, designed analyses, designed study, supervised analyses, supervised work, wrote paper, and obtained funding.

## Acknowledgements

We thank all study participants and their families as well as many involved institutions and their staff. This work was supported by grants from the National Institutes of Aging (R00AG075238, M.E.B.; R00AG070109, S.J.A.). This work was supported by access to equipment made possible by the Departments of Neurology at Washington University School of Medicine. We would like to thank the following resources for allowing access to their data: ADGC, ADSP, UK Biobank, and FinnGen. UK Biobank data were analyzed under Application Number 45420. Detailed acknowledgements for different genetic cohorts and biobanks are provided in the supplementary material. The FinnGen author list and contact information are provided in **Table-S9**.

