## SupplementFigures for "Shared Risk Genes and Casual Relationships across Sex Hormone Related Traits and Alzheimer’s Disease"

**Yang et al., 2026.**

### Supplemental Content

### Supplement Methods

##### Pleiotropy: Sex-stratified approach.

For pleiotropy analyses, we focused on sex-matched pairs: female Alzheimer's disease (AD) vs female hormone traits and male AD vs male hormone traits. In analyses aiming to identify pleiotropic loci (not global pleiotropic enrichment), we also paired the non-sex-stratified AD meta-analysis with female and male sex hormone-related trait GWAS respectively. The reasoning for this approach was to increase statistical power to identify pleiotropic loci, while subsequent tiered prioritization of pleiotropic loci filtered down to those loci displaying sex-biased associations with AD risk.

##### Pleiotropy: Negative control trait - natural hair color.

As a negative control trait, we additionally analyzed hair color phenotypes from UK Biobank genome-wide association summary statistics (UK Biobank GWAS Imputed v3, File Manifest Release 20180731)[1]. We considered five natural hair color categories assessed before greying—black, blonde, light brown, dark brown, and other. The red hair category was not included because of its low prevalence and because melanocortin-1 receptor (MC1R) variants underlying red hair have been implicated in neurodegenerative disease susceptibility, making red hair an uncertain negative control[2,3] .

##### Pleiotropy: Exclusion of complex LD regions and quality control.

To avoid linkage disequilibrium (LD)–related inflation of pleiotropic findings, GWAS variants were restricted to those falling outside of the major histocompatibility complex (6:28,510,120–33,480,577), the chromosome 8 inversion region (8:6,942,337–12,627,630), and the MAPT region (17:44,894,527–47,028,334). Variants were further intersected with European-ancestry samples from the 1000 Genomes Project to harmonize allele frequencies and LD structure, and LD-independent loci were extracted among variants surpassing the conjFDR significance threshold[4,5].

##### Genetic colocalization analyses.

To identify Tier-1 pleiotropic loci, we performed genetic colocalization (COLOC) analyses across sex hormone-related traits and AD. For these Tier-1 loci, we additionally performed COLOC analyses across the AD GWAS and QTL datasets. In either case, variants' genetic association signals were extracted for AD and each sex hormone-related GWAS or QTL dataset within a symmetric window centered on a respective locus' lead variant ($\pm$1Mb). Colocalization analyses were performed using the *coloc* (v.4.2.1) R package, applying coloc.abf under the standard single causal variant assumption. Standard prior probabilities were used (p1=1×10^-4, p2=1×10^-4, p12=1×10^-5). To relax this assumption and enable evaluation of multiple independent signals, we additionally used coloc.susie[6,7]. We considered two signals to have strong evidence of colocalization when the posterior probability of sharing a causal variant, PP4, was ≥0.7. For coloc.susie analyses, LD was estimated using an in-sample LD reference panel. We additionally performed visual quality control of colocalization results for Tier-1 loci using locuszoom plots. For three loci with low PP4 under coloc.abf but high PP4 under coloc.susie, we confirmed that the coloc.susie findings reflected false positives. These were excluded from the original set of 15 Tier-1 loci, retaining only 12 Tier-1 loci.

**Transcription factor motif enrichment analyses.**

Transcription factor motif enrichment was assessed using HOMER (findMotifs.pl) with the HOCOMOCO v11 motif collection[8,9]. Promoter regions were defined as −1000 to +300 bp around the annotated transcription start site of each gene. Two target gene sets were analyzed separately: genes with at least one AD–xQTL colocalization at female-biased Tier-1 loci and genes with at least one AD–xQTL colocalization at male-biased Tier-1 loci. As background, we used promoter regions of all GENCODE v48 protein-coding genes.

### Supplement Figures

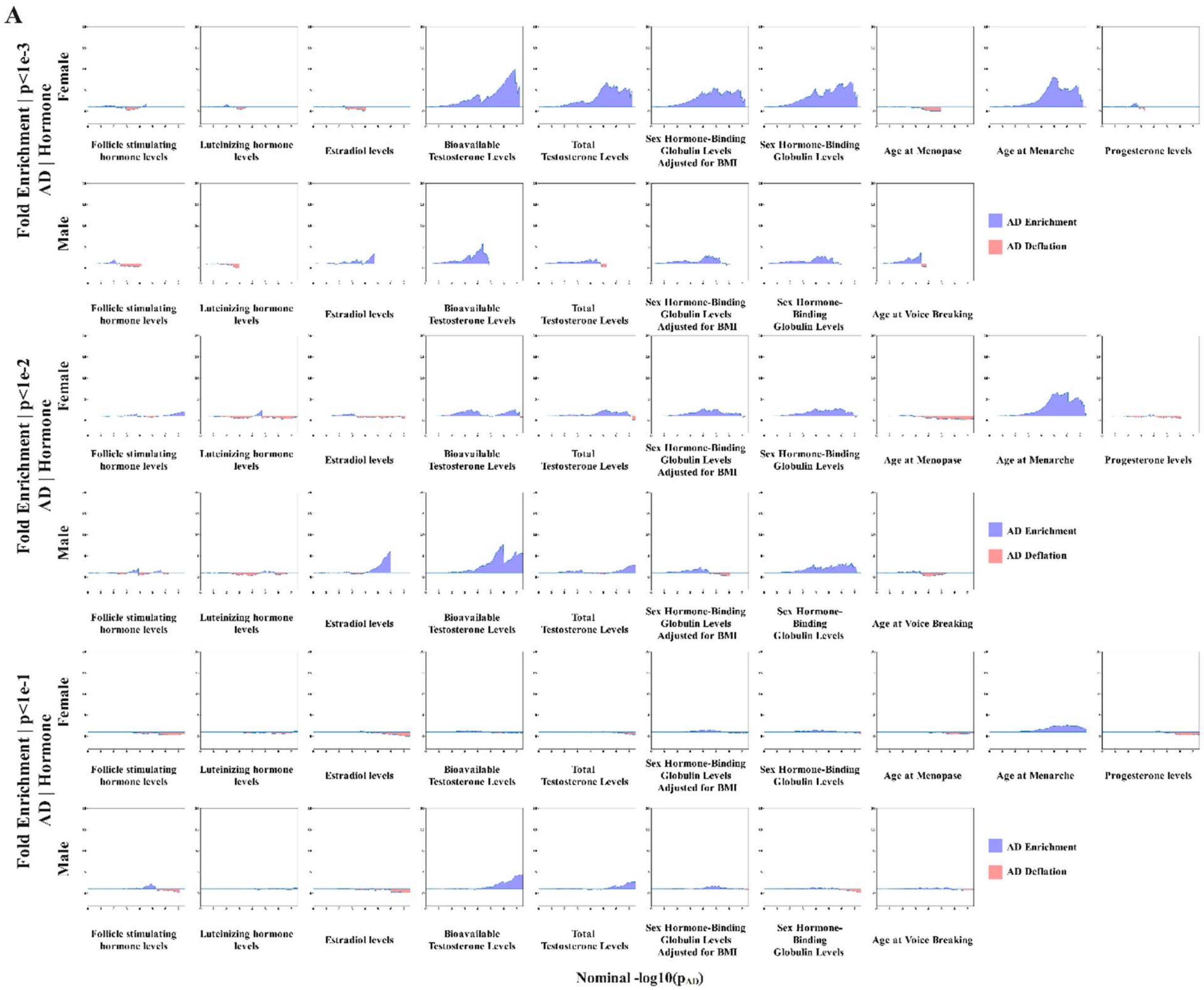

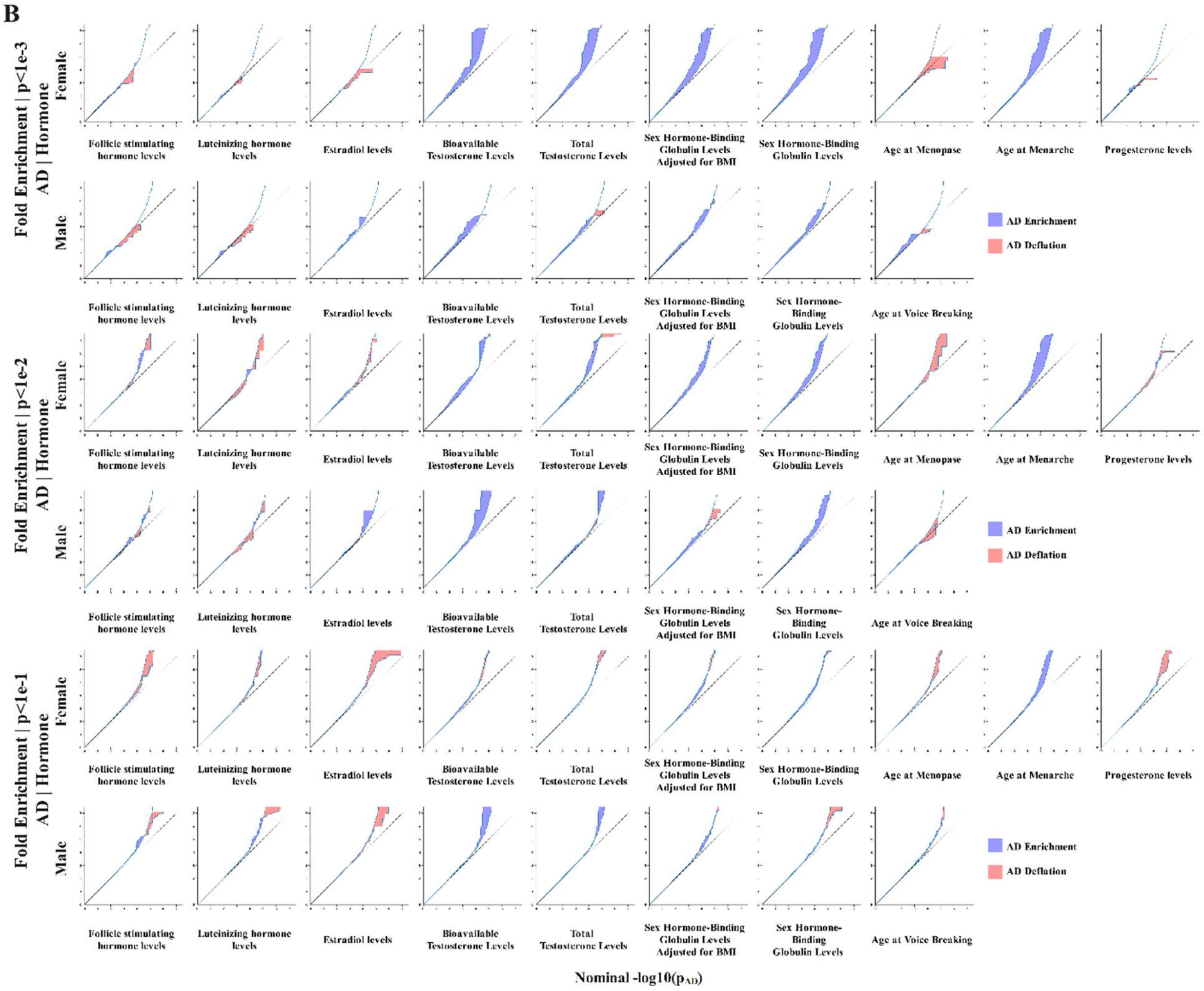

**Supplement Figure S1. AD fold-enrichment plots and Quantile-Quantile (QQ) plots conditioned on sex hormone-related traits.**

**A)** Sex-stratified AD fold-enrichment conditioned on sex hormone-related traits, shown at thresholds P < $10^{-3}$, $10^{-2}$, $10^{-1}$ (rows) across traits (columns) for females and males. The horizontal baseline denotes the null (fold enrichment = 1). **B)** QQ plots of nominal -$\log_{10}(P_{(AD)})$ versus empirical -$\log_{10}(q_{(AD)})$ for the same thresholds and strata as in (A). **A-B)** Blue shaded areas indicate enrichment while red indicate deflation, when conditioning variants on their associations with sex hormone-related traits.

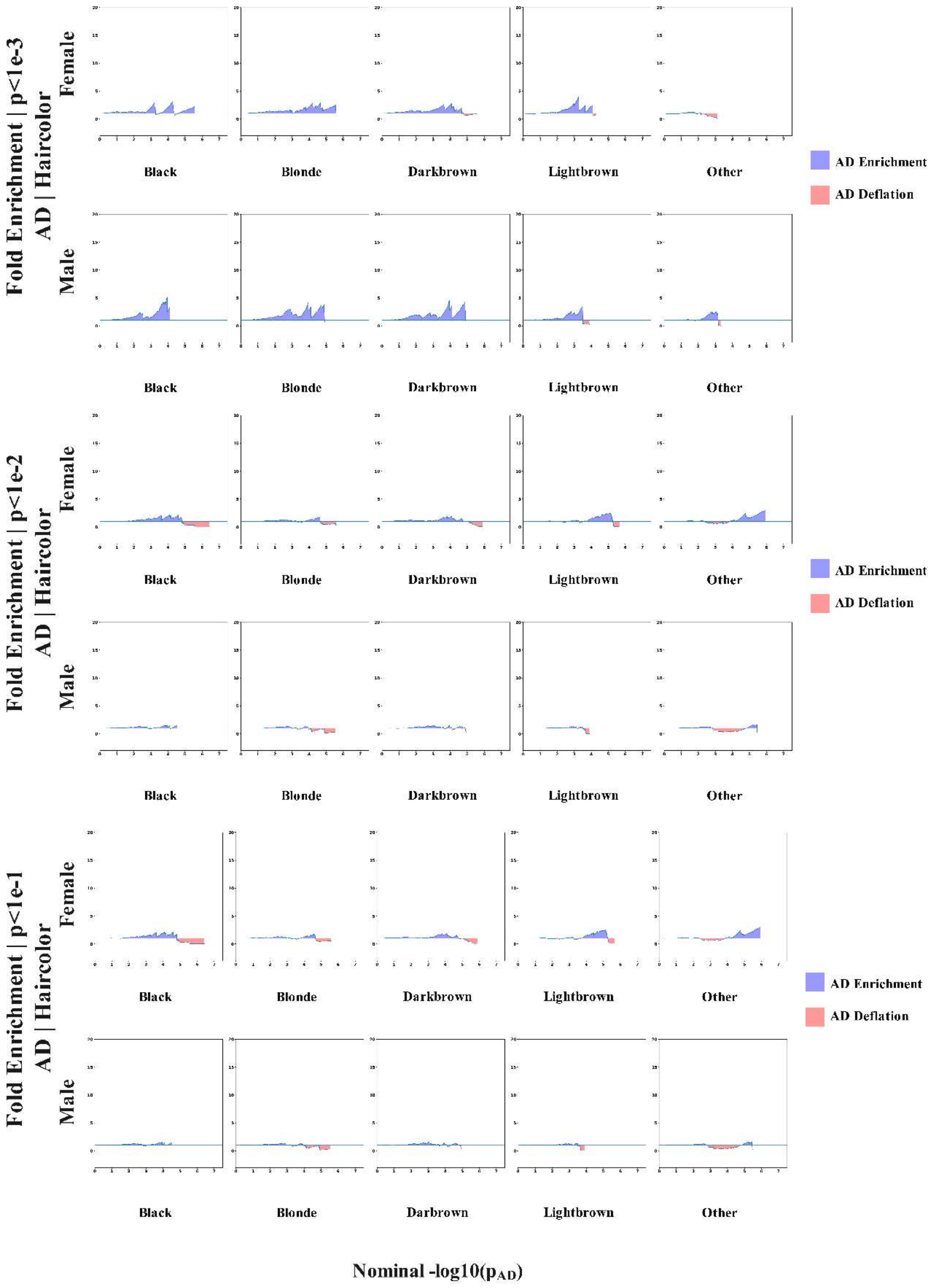
Fold Enrichment | p<1e-3
AD | Haircolor
Female
Male
Black
Blonde
Darkbrown
Lightbrown
Other
AD Enrichment
AD Deflation
Fold Enrichment | p<1e-2
AD | Haircolor
Female
Male
Black
Blonde
Darkbrown
Lightbrown
Other
AD Enrichment
AD Deflation
Fold Enrichment | p<1e-1
AD | Haircolor
Female
Male
Black
Blonde
Darkbrown
Lightbrown
Other
AD Enrichment
AD Deflation
Black
Blonde
Darbrown
Lightbrown
Other
Nominal -log10($p_{AD}$)

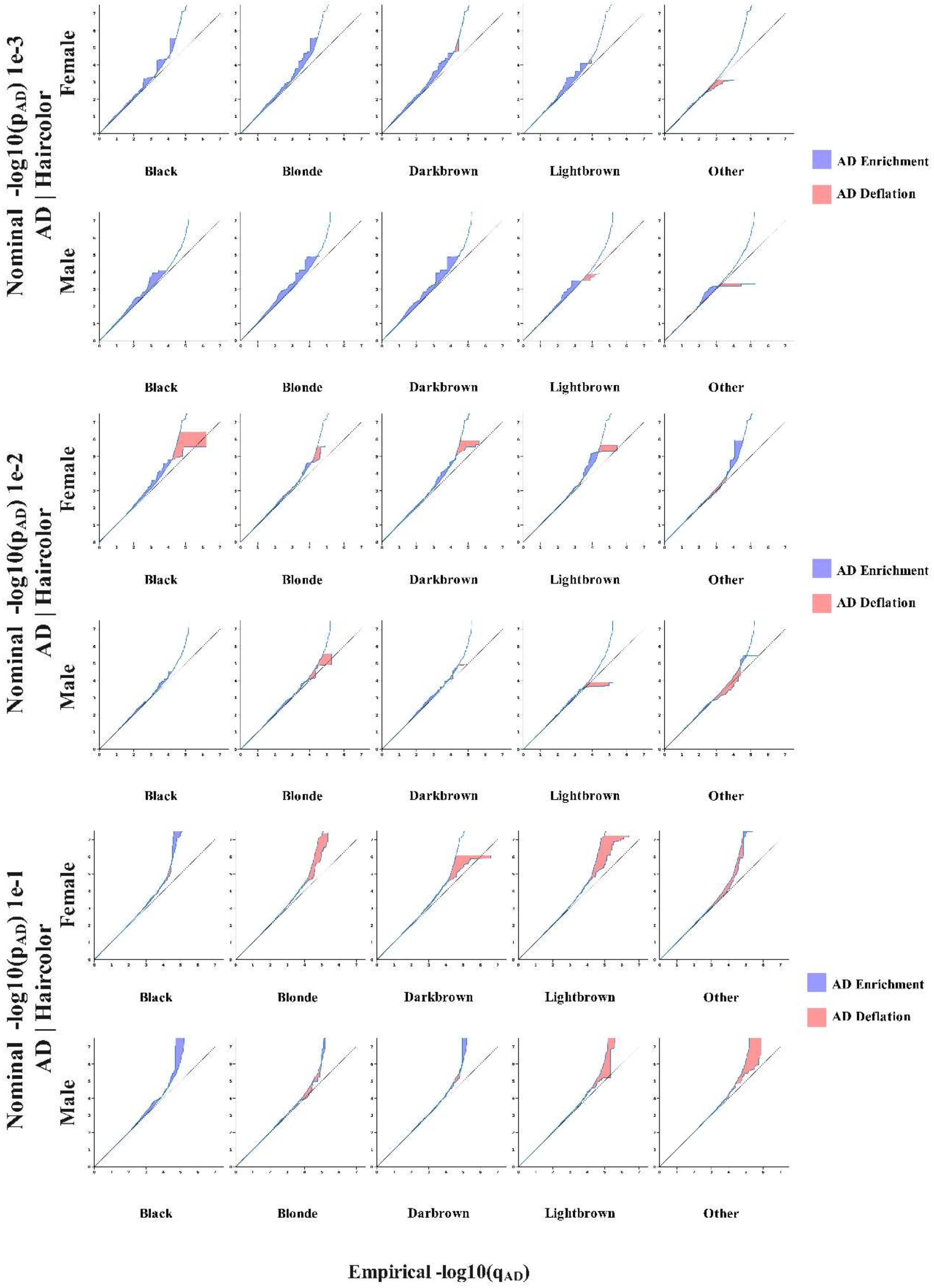
Nominal -log10(p_AD) 1e-3
AD | Haircolor
Female
Male
Black
Blonde
Darkbrown
Lightbrown
Other
AD Enrichment
AD Deflation
Black
Blonde
Darkbrown
Lightbrown
Other
Nominal -log10(p_AD) 1e-2
AD | Haircolor
Female
Male
Black
Blonde
Darkbrown
Lightbrown
Other
AD Enrichment
AD Deflation
Black
Blonde
Darkbrown
Lightbrown
Other
Nominal -log10(p_AD) 1e-1
AD | Haircolor
Female
Male
Black
Blonde
Darkbrown
Lightbrown
Other
AD Enrichment
AD Deflation
Black
Blonde
Darbrown
Lightbrown
Other
Empirical -log10(q_AD)

**Supplement Figure S2. AD fold-enrichment plots and Quantile-Quantile (QQ) plots conditioned on natural hair color as negative control.**

**A)** Sex-stratified AD fold-enrichment conditioned on natural hair color traits, shown at thresholds $P < 10^{-3}$, $10^{-2}$, $10^{-1}$ (rows) across traits (columns) for females and males. The horizontal baseline denotes the null (fold enrichment = 1). **B)** QQ plots of nominal $-\log_{10}(P_{(AD)})$ versus empirical $-\log_{10}(q_{(AD)})$ for the same thresholds and strata as in (A). **A-B)** Blue shaded areas indicate enrichment while red indicate deflation, when conditioning variants on their associations with natural hair color traits. Note that the x-axis and y-axis limits are the same as those in eFigure-1 to enable direct comparison.

**Alzheimer's Disease by Total Testosterone Female**

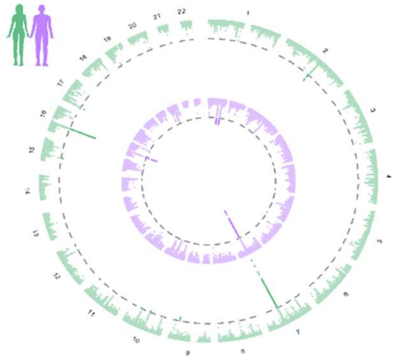

**Alzheimer's Disease by Bioavailable Testosterone Female**

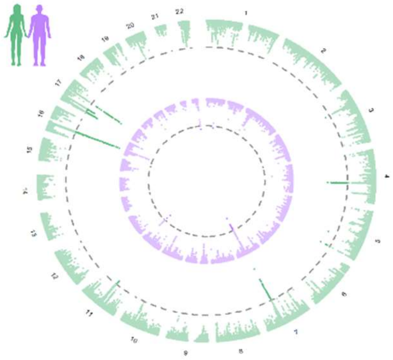

**Alzheimer's Disease by SHBG**

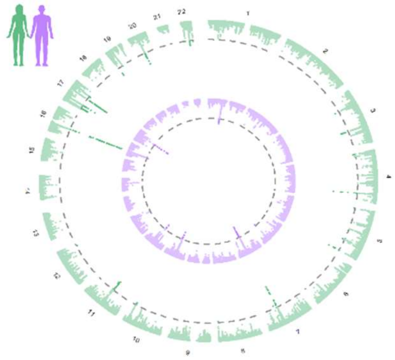

**Alzheimer's Disease by SHBG adjusted by BMI**

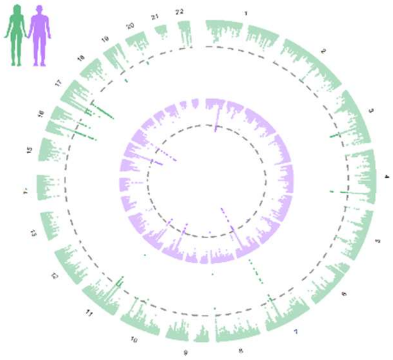

**Alzheimer's Disease by Age at Menarche**

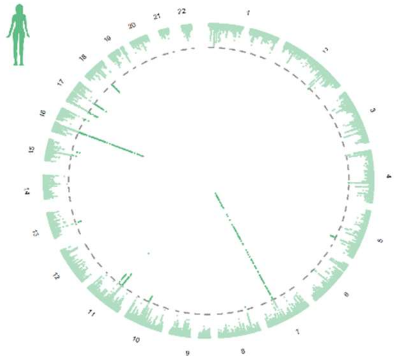

**Alzheimer's Disease by Age at Menopause**

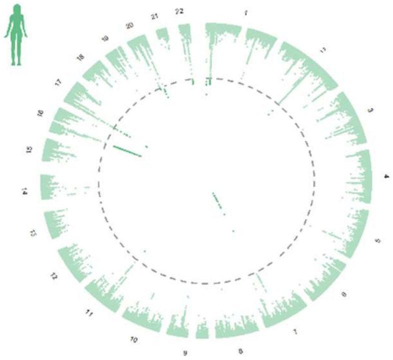

**Alzheimer's Disease by Age at voicebreaking**

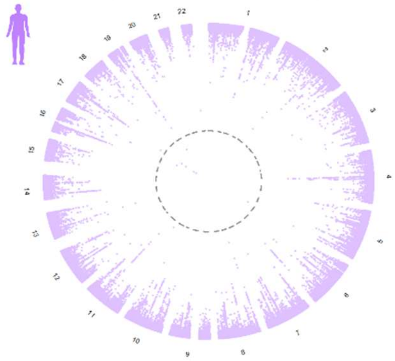

**Supplement Figure S3. Circos plots of genome-wide pleiotropy between AD and sex hormone-related traits.**

Panels show AD by: total testosterone, bioavailable testosterone, SHBG, SHBG adjusted for BMI, age at menarche, age at menopause, and age at voice breaking. Outer ring (green): female results; inner ring (purple): male results. The dotted circle marks the significance threshold ($P_{FDR} < 0.05$).

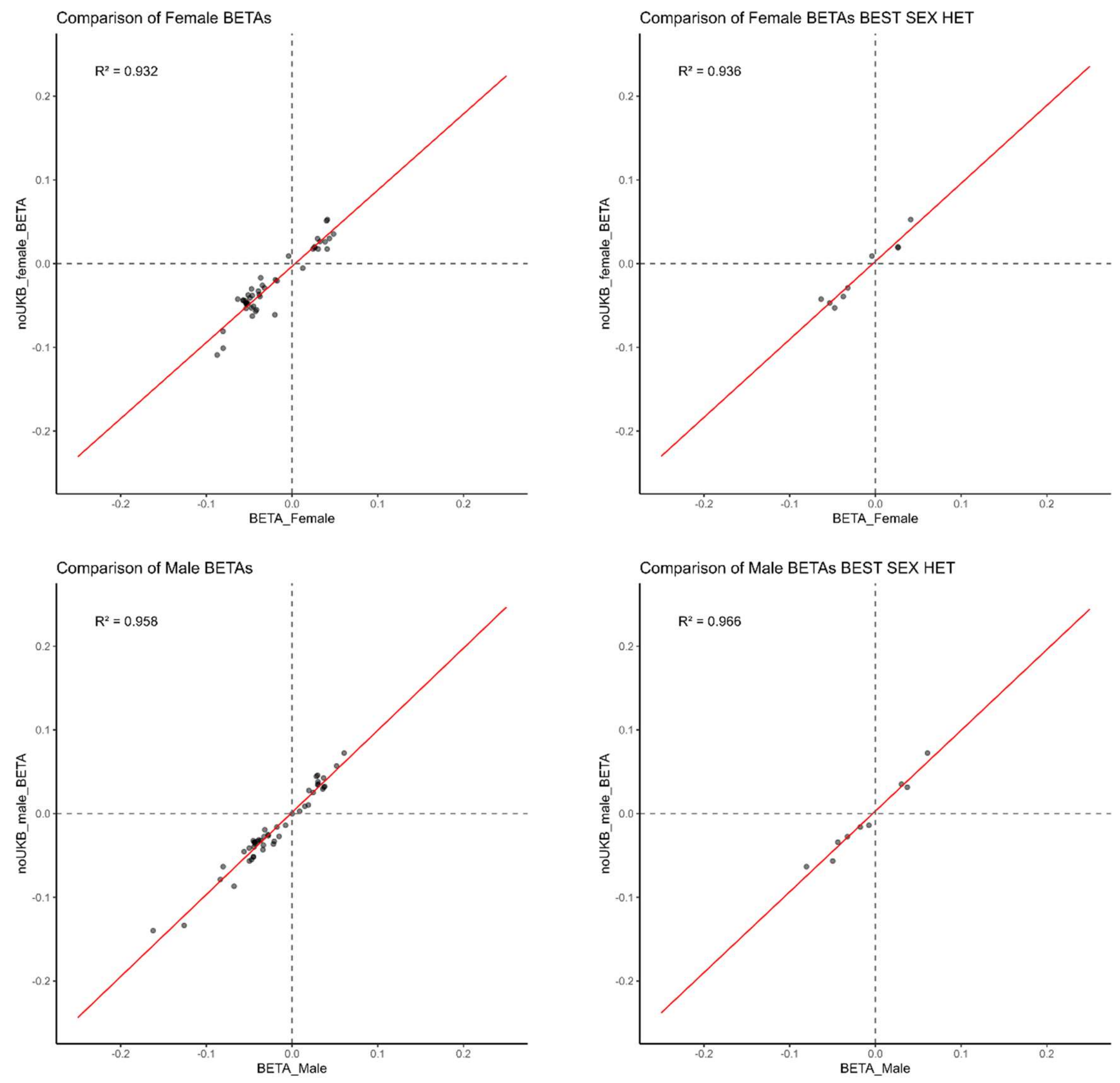

**Supplement Figure S4. Comparison of tier 1 prioritized SNPs' effect sizes (β) from sex-stratified AD GWAS with versus without UKB samples.**

Left: all Tier-1 index SNPs. Right: LD-clumped subset retaining the SNP with the strongest sex-heterogeneity signal per Tier-1 locus. Each point represents a SNP; the red line is a linear fit; dashed lines denote β = 0. Reported squared Pearson correlation $R^2$ values summarize the overall similarity of effect sizes between the two analyses.

**FigureS5.1**

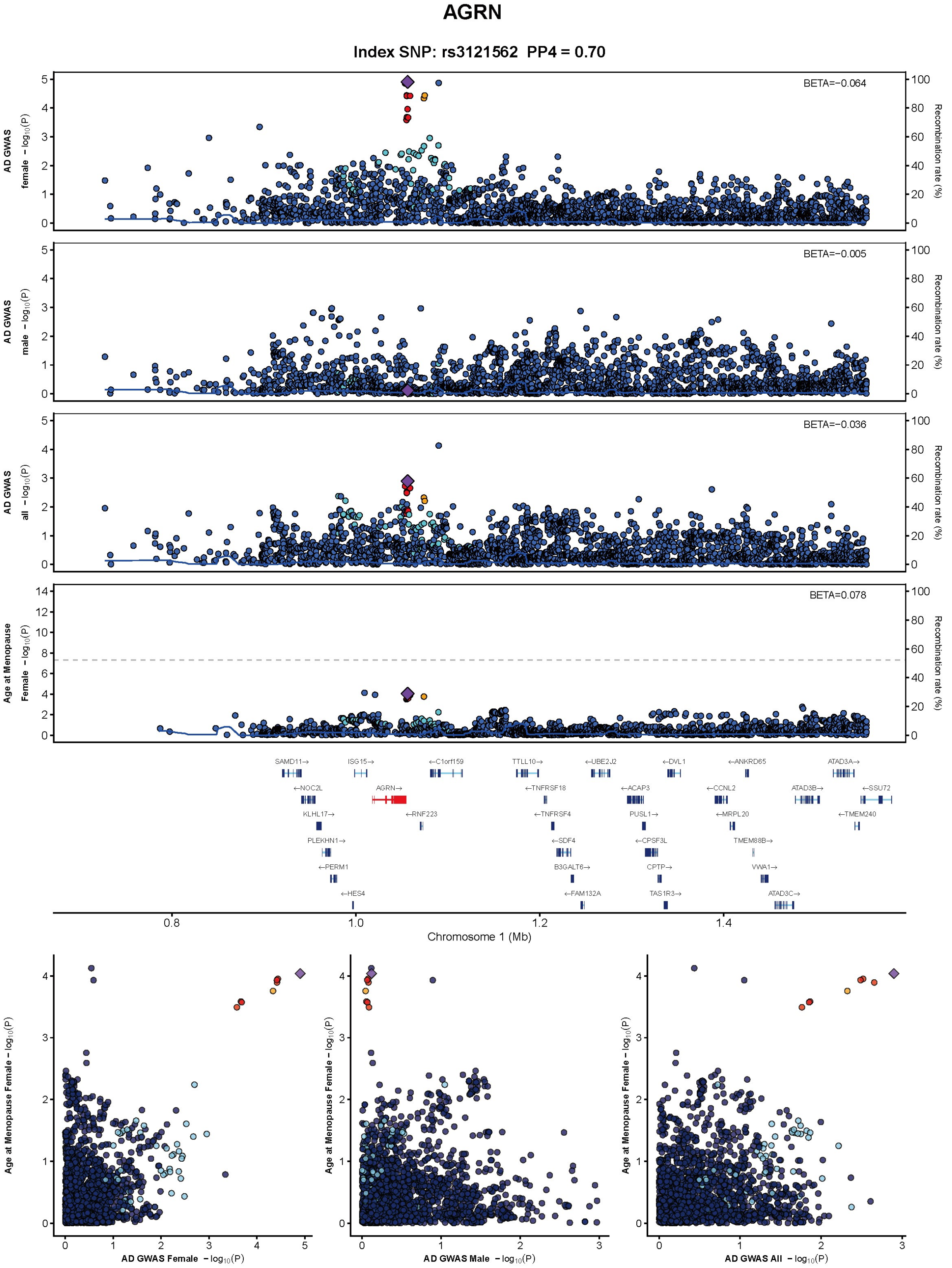
AGRN
Index SNP: rs3121562 PP4 = 0.70
AD GWAS female − log10(P)
BETA=−0.064
AD GWAS male − log10(P)
BETA=−0.005
AD GWAS all − log10(P)
BETA=−0.036
Age at Menopause Female − log10(P)
BETA=0.078
Recombination rate (%)
SAMD11→
ISG15→
←C1orf159
TTLL10→
←UBE2J2
←DVL1
←ANKRD65
ATAD3A→
←NOC2L
AGRN→
←TNFRSF18
←ACAP3
←CCNL2
ATAD3B→
←SSU72
KLHL17→
←RNF223
←TNFRSF4
PUSL1→
←MRPL20
←TMEM240
PLEKHN1→
←SDF4
←CPSF3L
TMEM88B→
←PERM1
B3GALT6→
CPTP→
VWA1→
←HES4
←FAM132A
TAS1R3→
ATAD3C→
Chromosome 1 (Mb)
Age at Menopause Female − log10(P)
AD GWAS Female − log10(P)
AD GWAS Male − log10(P)
AD GWAS All − log10(P)

**FigureS5.2**

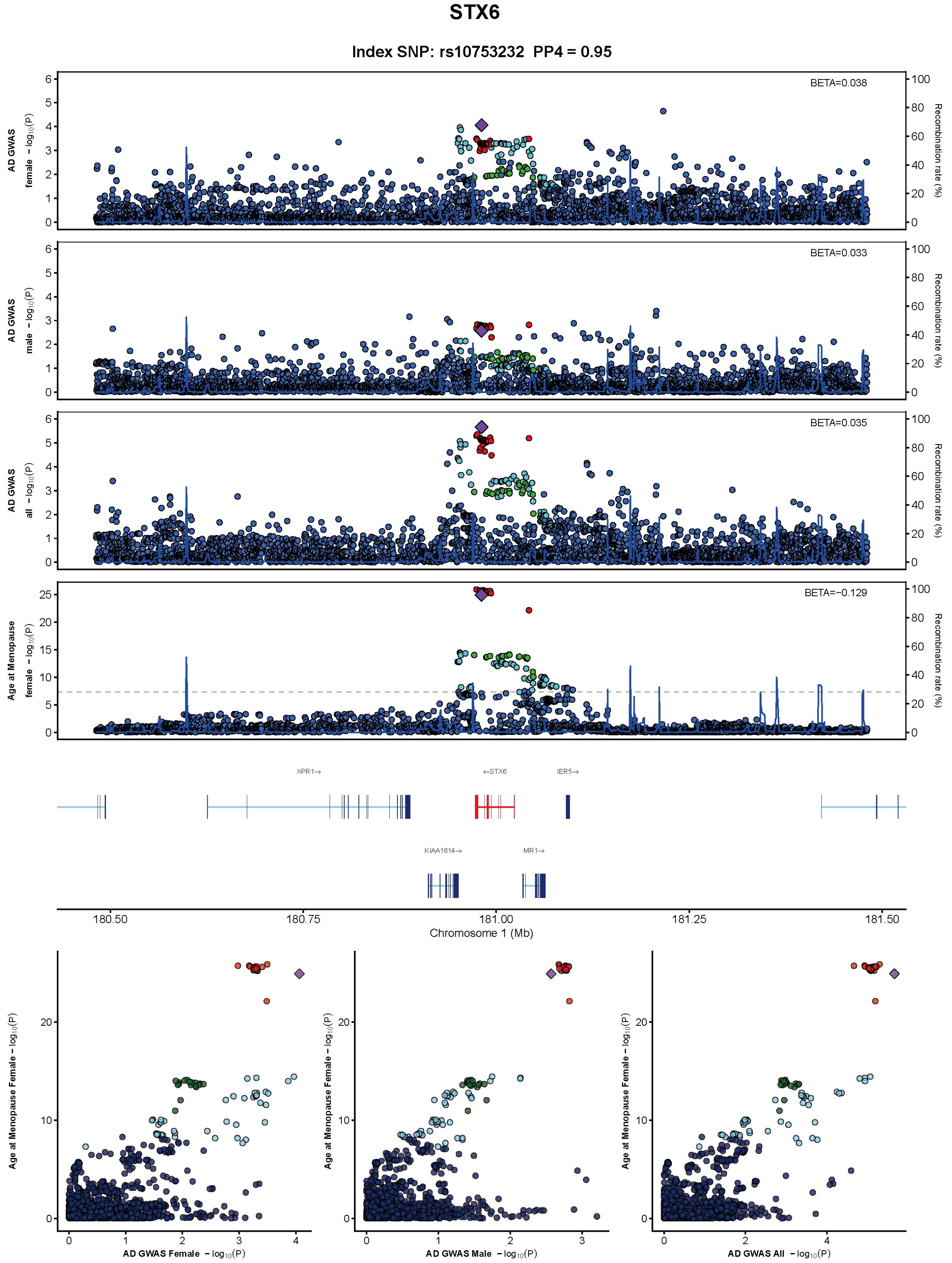
STX6
Index SNP: rs10753232 PP4 = 0.95
BETA=0.038
BETA=0.033
BETA=0.035
BETA=−0.129
AD GWAS female − log10(P)
AD GWAS male − log10(P)
AD GWAS all − log10(P)
Age at Menopause female − log10(P)
Recombination rate (%)
XPR1→
←STX6
IER5→
KIAA1614→
MR1→
180.50
180.75
181.00
181.25
181.50
Chromosome 1 (Mb)
Age at Menopause Female − log10(P)
AD GWAS Female − log10(P)
AD GWAS Male − log10(P)
AD GWAS All − log10(P)

**FigureS5.3**

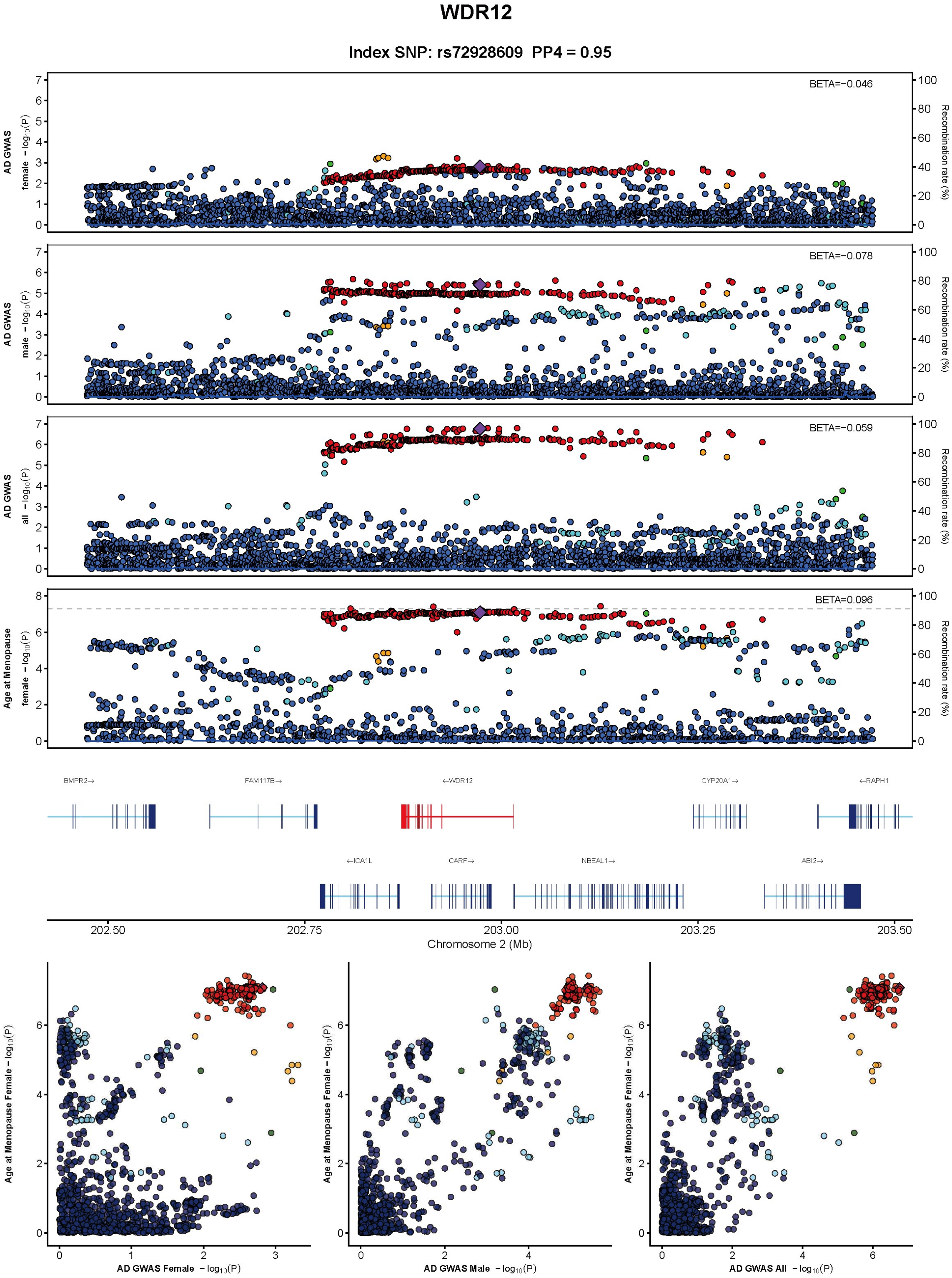
WDR12
Index SNP: rs72928609 PP4 = 0.95
BETA=−0.046
BETA=−0.078
BETA=−0.059
BETA=0.096
AD GWAS female − log10(P)
AD GWAS male − log10(P)
AD GWAS all − log10(P)
Age at Menopause female − log10(P)
Recombination rate (%)
BMPR2→
FAM117B→
←WDR12
CYP20A1→
←RAPH1
←ICA1L
CARF→
NBEAL1→
ABI2→
Chromosome 2 (Mb)
202.50
202.75
203.00
203.25
203.50
Age at Menopause Female − log10(P)
AD GWAS Female − log10(P)
AD GWAS Male − log10(P)
AD GWAS All − log10(P)

**FigureS5.4**

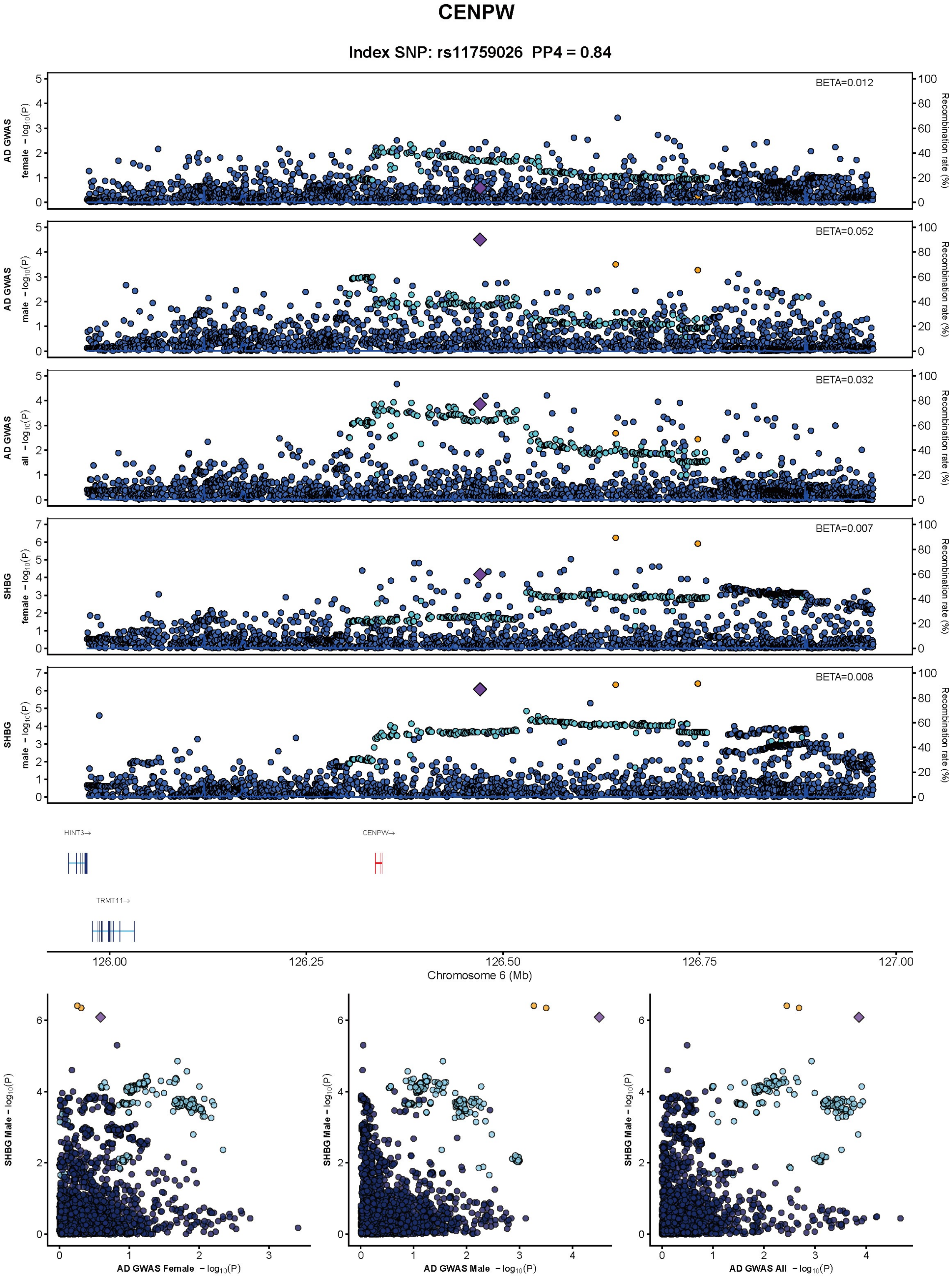

CENPW
Index SNP: rs11759026 PP4 = 0.84
BETA=0.012
BETA=0.052
BETA=0.032
BETA=0.007
BETA=0.008
AD GWAS female − log10(P)
AD GWAS male − log10(P)
AD GWAS all − log10(P)
SHBG female − log10(P)
SHBG male − log10(P)
Recombination rate (%)
HINT3→
CENPW→
TRMT11→
126.00
126.25
126.50
126.75
127.00
Chromosome 6 (Mb)
SHBG Male − log10(P)
AD GWAS Female − log10(P)
AD GWAS Male − log10(P)
AD GWAS All − log10(P)

**FigureS5.5**

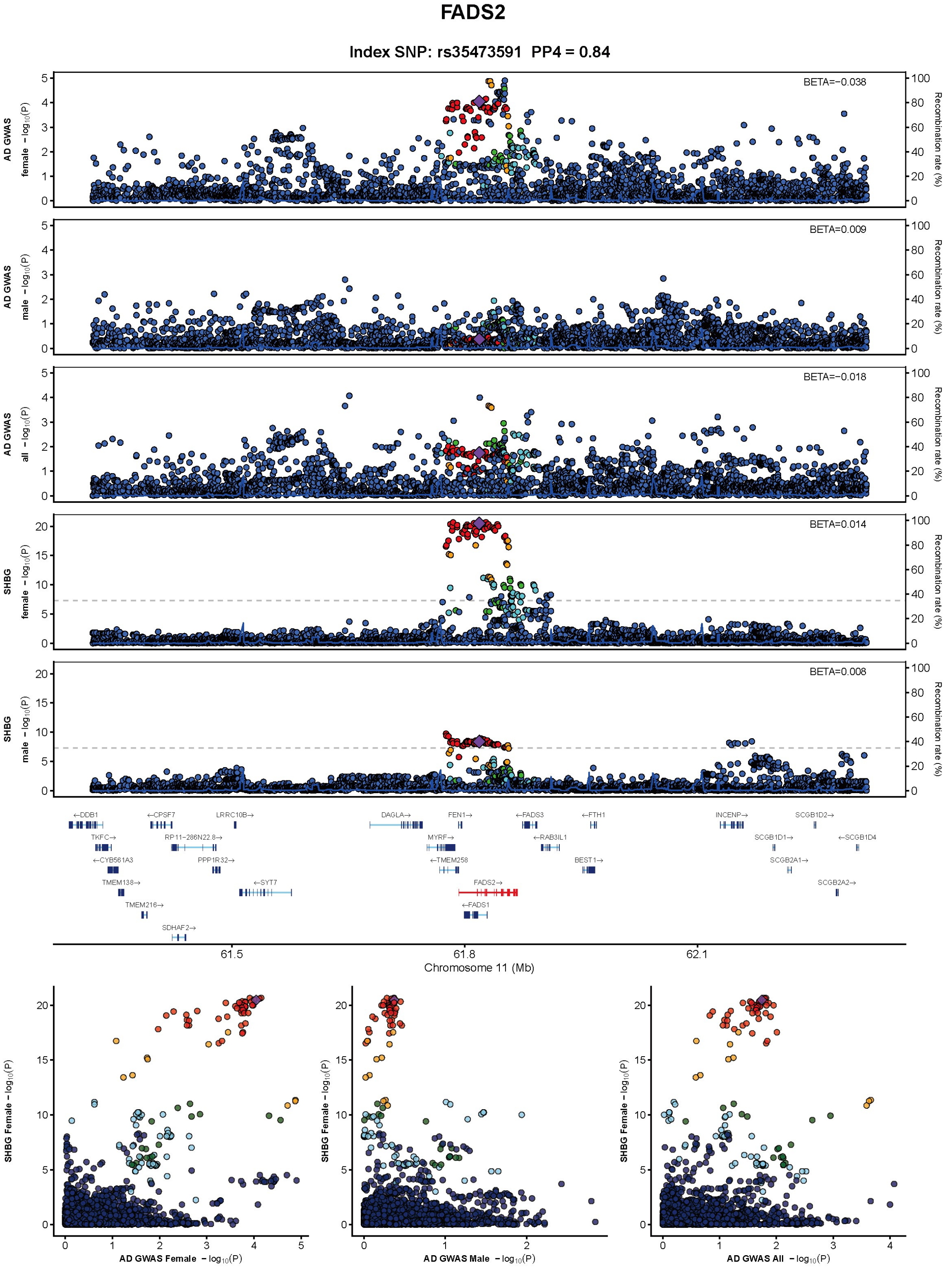
FADS2
Index SNP: rs35473591 PP4 = 0.84
AD GWAS female −log10(P)
BETA=−0.038
AD GWAS male −log10(P)
BETA=0.009
AD GWAS all −log10(P)
BETA=−0.018
SHBG female −log10(P)
BETA=0.014
SHBG male −log10(P)
BETA=0.008
Recombination rate (%)
61.5
61.8
62.1
Chromosome 11 (Mb)
SHBG Female −log10(P)
AD GWAS Female −log10(P)
AD GWAS Male −log10(P)
AD GWAS All −log10(P)

**FigureS5.6**

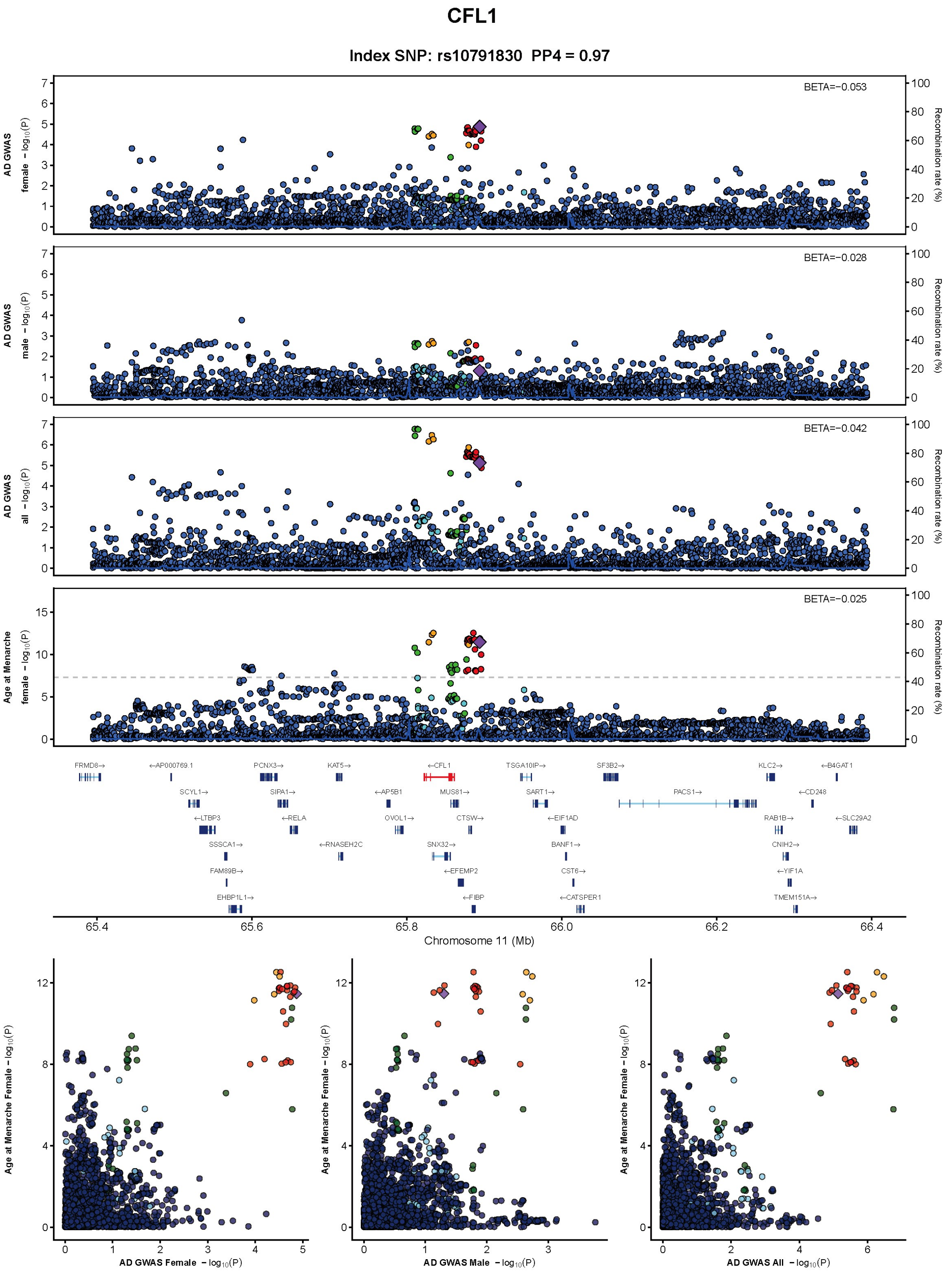
CFL1
Index SNP: rs10791830 PP4 = 0.97
BETA=−0.053
BETA=−0.028
BETA=−0.042
BETA=−0.025
AD GWAS female −log10(P)
AD GWAS male −log10(P)
AD GWAS all −log10(P)
Age at Menarche female −log10(P)
Recombination rate (%)
Chromosome 11 (Mb)
AD GWAS Female −log10(P)
AD GWAS Male −log10(P)
AD GWAS All −log10(P)
Age at Menarche Female −log10(P)

**FigureS5.7**

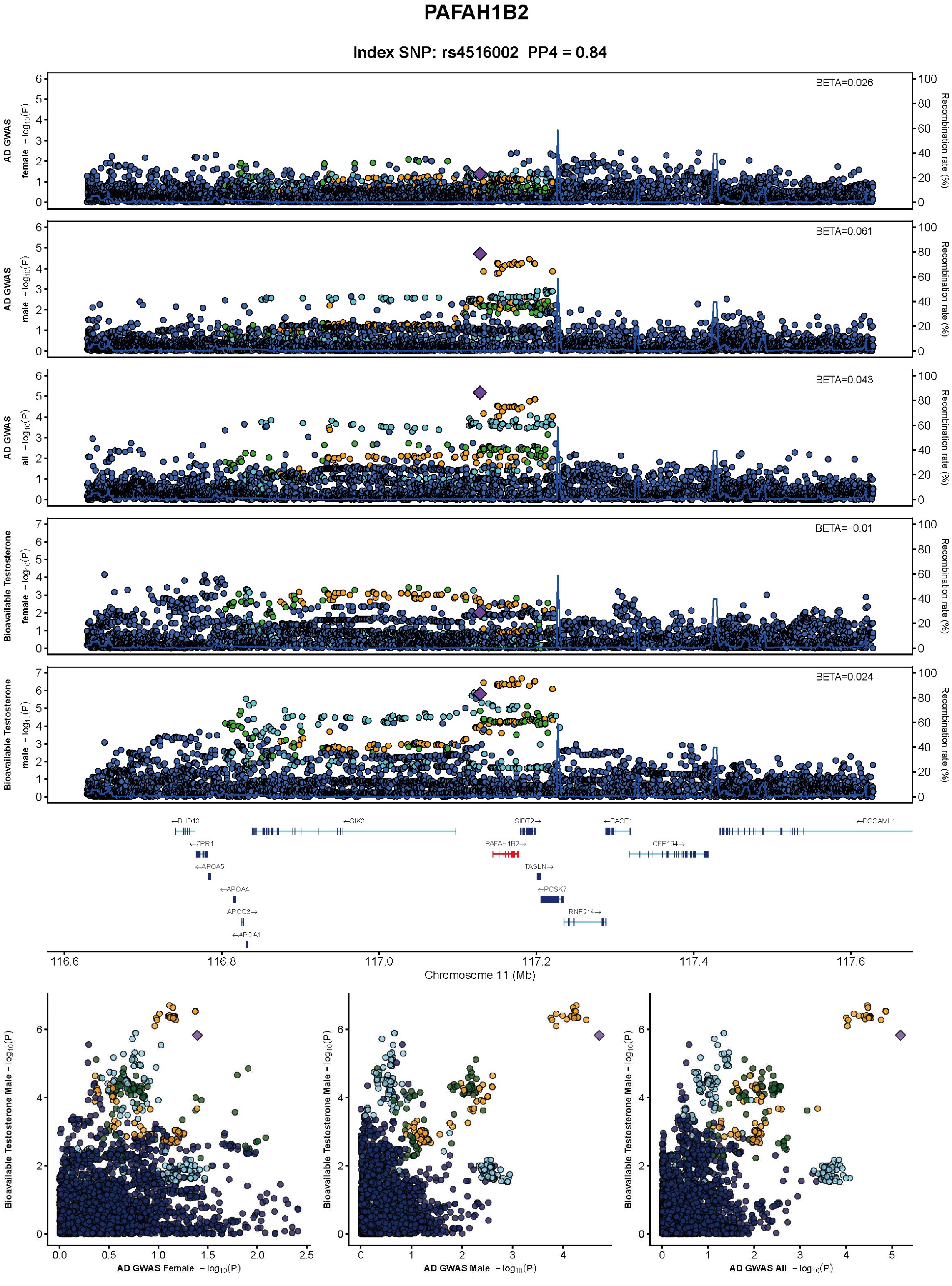
PAFAH1B2
Index SNP: rs4516002 PP4 = 0.84
BETA=0.026
BETA=0.061
BETA=0.043
BETA=−0.01
BETA=0.024
AD GWAS female −log10(P)
AD GWAS male −log10(P)
AD GWAS all −log10(P)
Bioavailable Testosterone female −log10(P)
Bioavailable Testosterone male −log10(P)
Recombination rate (%)
←BUD13
←SIK3
SIDT2→
←BACE1
←DSCAML1
←ZPR1
PAFAH1B2→
CEP164→
←APOA5
TAGLN→
←APOA4
←PCSK7
APOC3→
RNF214→
←APOA1
116.6
116.8
117.0
117.2
117.4
117.6
Chromosome 11 (Mb)
Bioavailable Testosterone Male −log10(P)
AD GWAS Female −log10(P)
AD GWAS Male −log10(P)
AD GWAS All −log10(P)

**FigureS5.8**

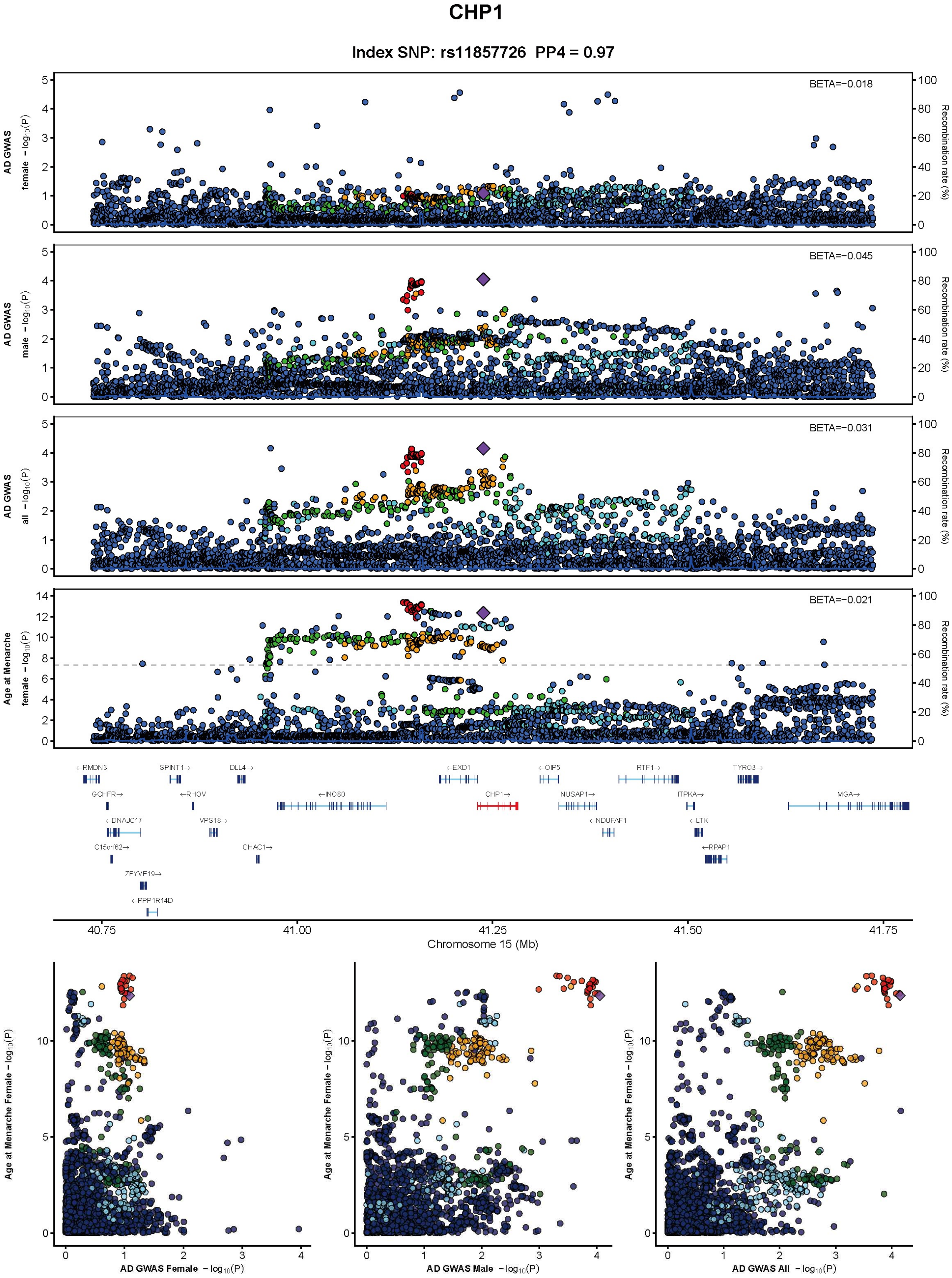

CHP1
Index SNP: rs11857726 PP4 = 0.97
BETA=−0.018
BETA=−0.045
BETA=−0.031
BETA=−0.021
AD GWAS female −log10(P)
AD GWAS male −log10(P)
AD GWAS all −log10(P)
Age at Menarche female −log10(P)
Recombination rate (%)
Chromosome 15 (Mb)
AD GWAS Female −log10(P)
AD GWAS Male −log10(P)
AD GWAS All −log10(P)
Age at Menarche Female −log10(P)

**FigureS5.9**

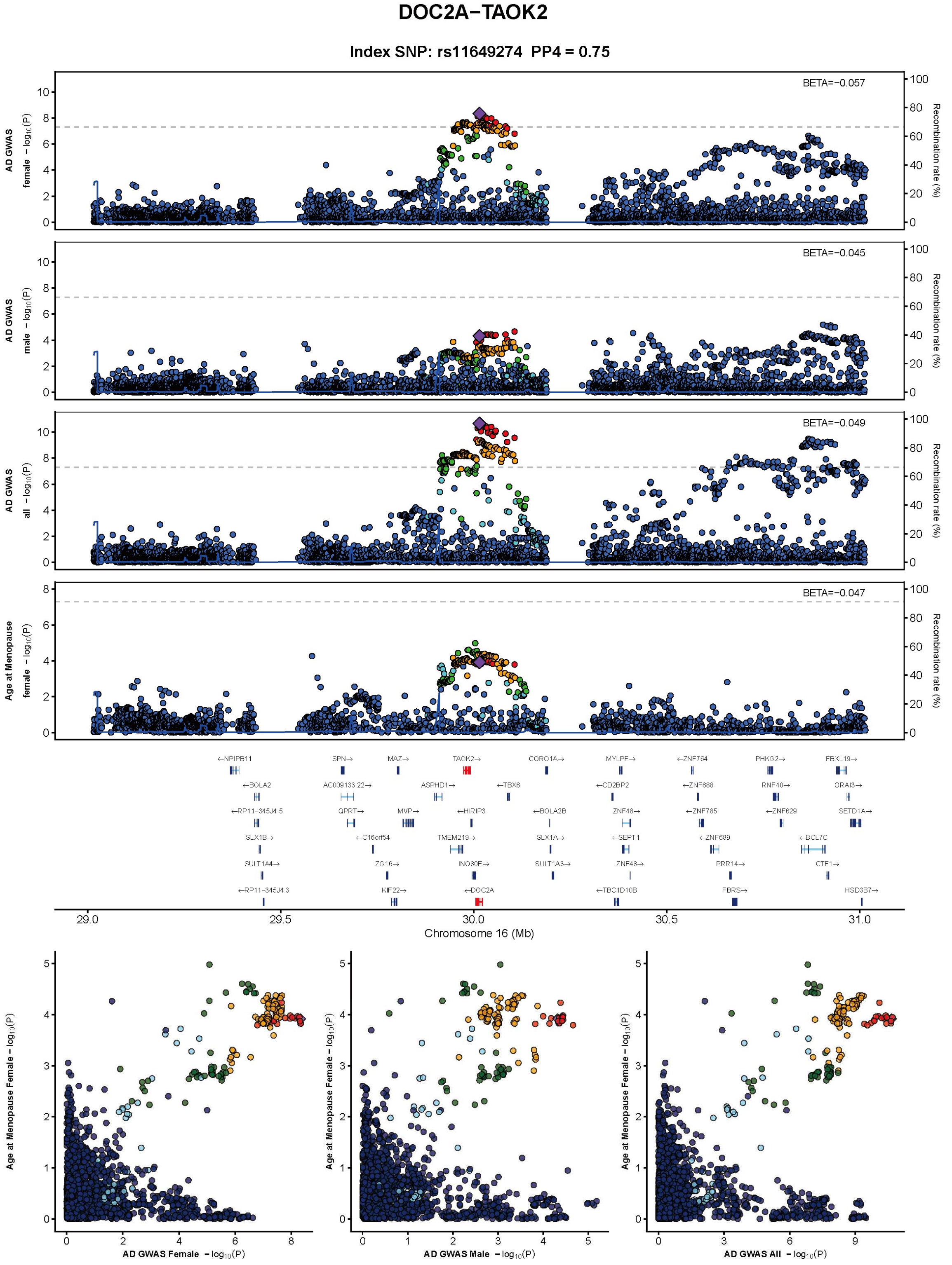
DOC2A−TAOK2
Index SNP: rs11649274 PP4 = 0.75
BETA=−0.057
BETA=−0.045
BETA=−0.049
BETA=−0.047
AD GWAS female −log10(P)
AD GWAS male −log10(P)
AD GWAS all −log10(P)
Age at Menopause female −log10(P)
Recombination rate (%)
Chromosome 16 (Mb)
AD GWAS Female −log10(P)
AD GWAS Male −log10(P)
AD GWAS All −log10(P)
Age at Menopause Female −log10(P)

**FigureS5.10**

#### WDR81

**Index SNP: rs11078597  PP4 = 0.95**

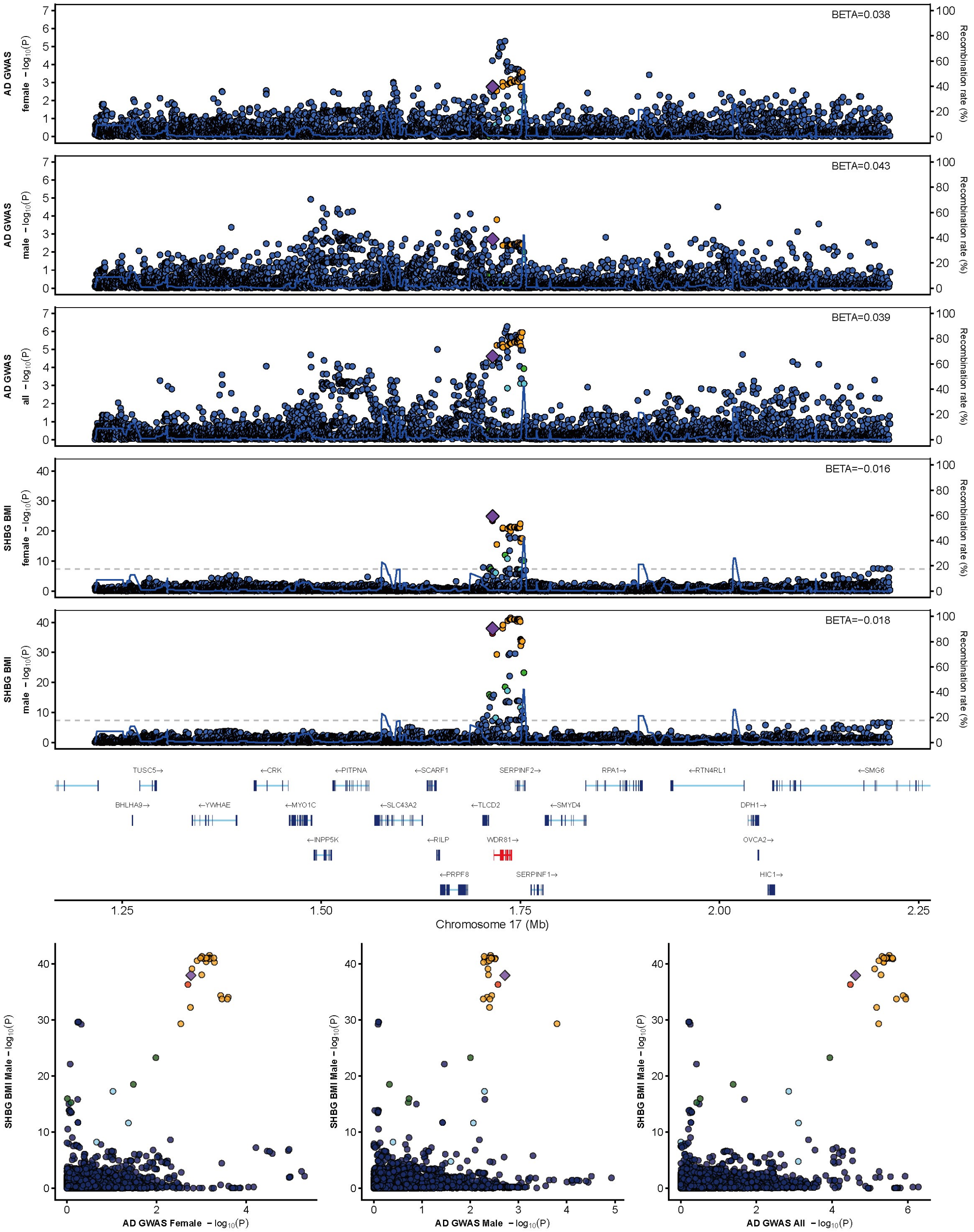

**FigureS5.11**

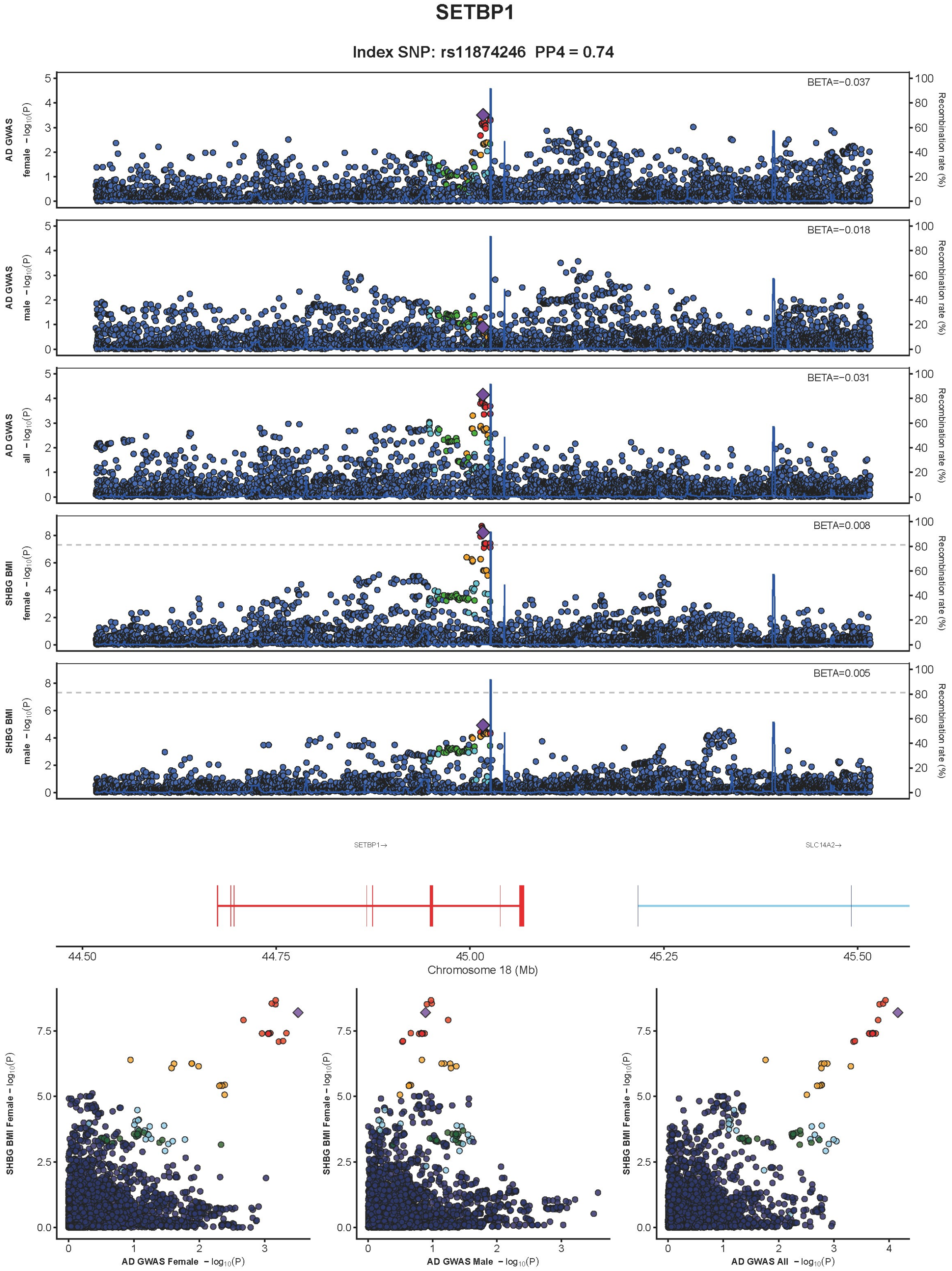

SETBP1
Index SNP: rs11874246 PP4 = 0.74
AD GWAS female − log10(P)
BETA=−0.037
AD GWAS male − log10(P)
BETA=−0.018
AD GWAS all − log10(P)
BETA=−0.031
SHBG BMI female − log10(P)
BETA=0.008
SHBG BMI male − log10(P)
BETA=0.005
Recombination rate (%)
SETBP1→
SLC14A2→
44.50
44.75
45.00
45.25
45.50
Chromosome 18 (Mb)
SHBG BMI Female − log10(P)
AD GWAS Female − log10(P)
AD GWAS Male − log10(P)
AD GWAS All − log10(P)

**FigureS5.12**

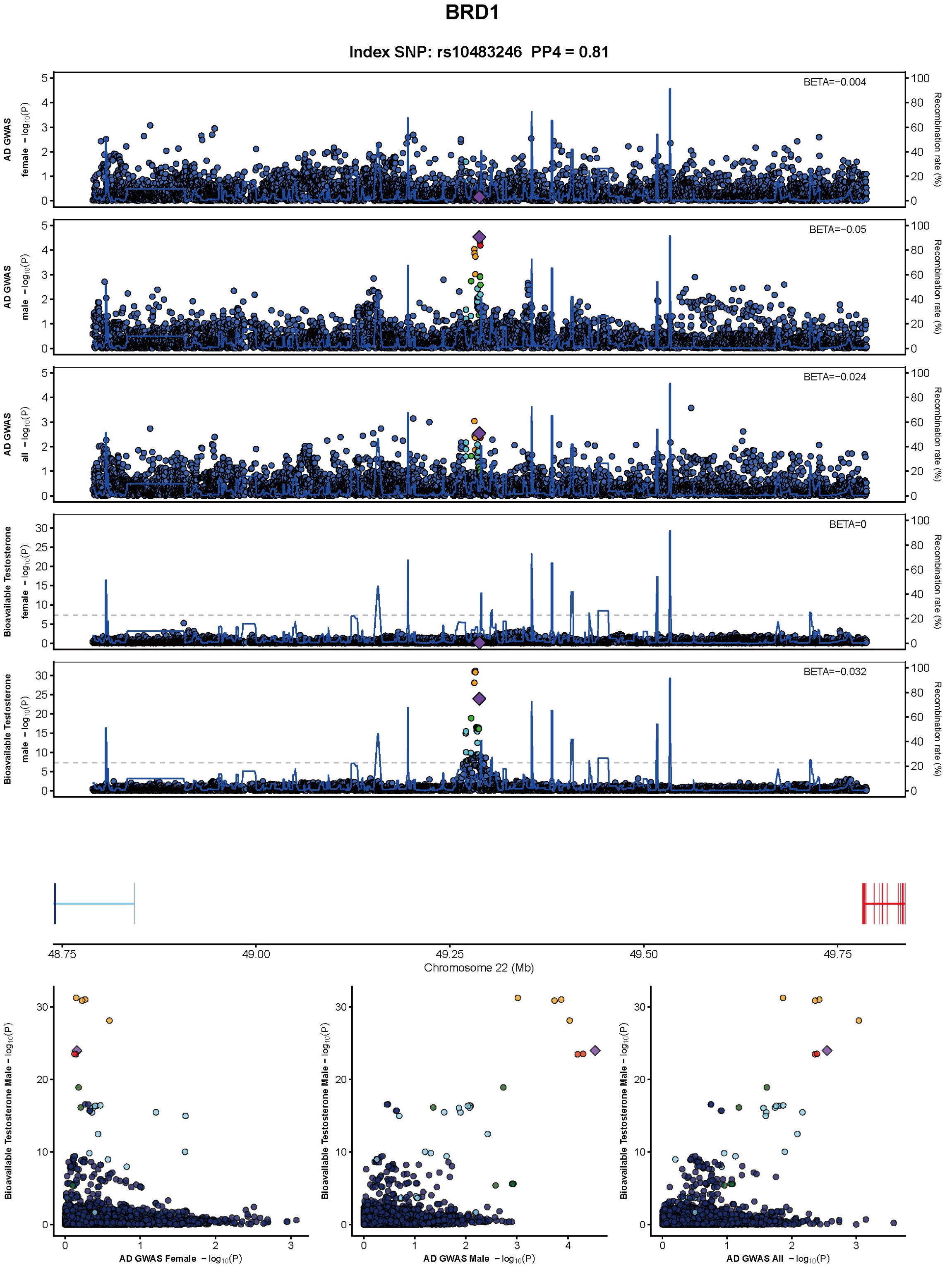
BRD1
Index SNP: rs10483246 PP4 = 0.81
AD GWAS female − log10(P)
AD GWAS male − log10(P)
AD GWAS all − log10(P)
Bioavailable Testosterone female − log10(P)
Bioavailable Testosterone male − log10(P)
Recombination rate (%)
BETA=−0.004
BETA=−0.05
BETA=−0.024
BETA=0
BETA=−0.032
48.75
49.00
49.25
49.50
49.75
Chromosome 22 (Mb)
Bioavailable Testosterone Male − log10(P)
AD GWAS Female − log10(P)
AD GWAS Male − log10(P)
AD GWAS All − log10(P)

**Supplement Figure S5. Detailed locus zoom and locus-compare plots at prioritized Tier-1 pleiotropic loci.**

For each Tier-1 locus, top panels show regional association plots for female, male, and sex-combined AD GWAS together with the corresponding sex hormone-related trait GWAS. Y-axes show $-\log_{10}(P)$, the purple diamond marks the index variant, and point colors indicate linkage disequilibrium with that variant. PP4 values denote support for a shared causal variant from genetic colocalization analyses. Bottom panels show locus-compare plots between AD and the corresponding sex hormone-related trait. Variants are colored by linkage disequilibrium ($r^2$) with the index variant: red ($r^2 \geqslant 0.8$), orange ($0.6 \leqslant r^2 < 0.8$), green ($0.4 \leqslant r^2 < 0.6$), light blue ($0.2 \leqslant r^2 < 0.4$), and dark blue ($r^2 < 0.2$).

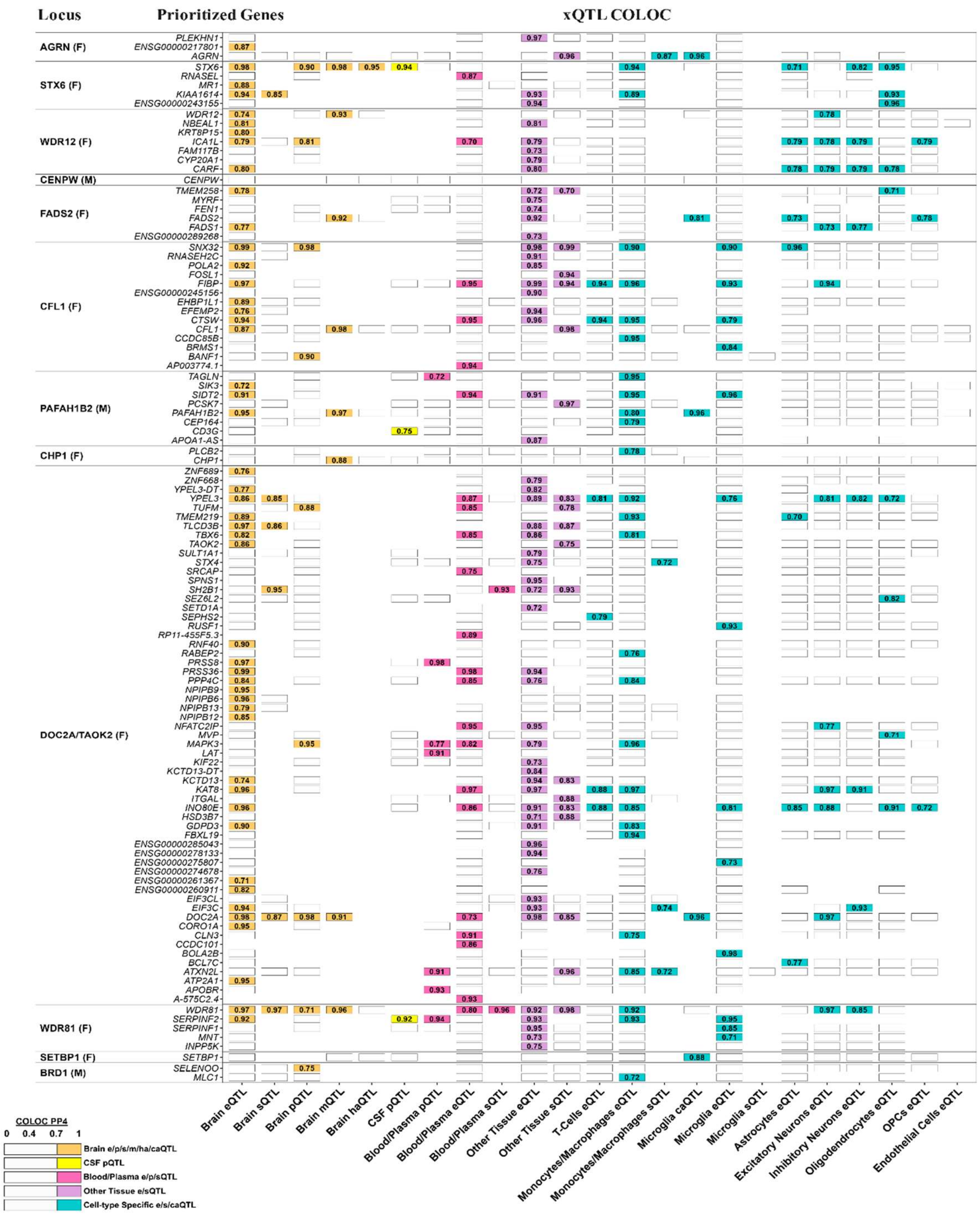

Locus
Prioritized Genes
xQTL COLOC
AGRN (F)
STX6 (F)
WDR12 (F)
CENPW (M)
FADS2 (F)
CFL1 (F)
PAFAH1B2 (M)
CHP1 (F)
DOC2A/TAOK2 (F)
WDR81 (F)
SETBP1 (F)
BRD1 (M)
Brain eQTL
Brain sQTL
Brain pQTL
Brain mQTL
Brain haQTL
CSF pQTL
Blood/Plasma pQTL
Blood/Plasma eQTL
Blood/Plasma sQTL
Other Tissue eQTL
Other Tissue sQTL
T-Cells eQTL
Monocytes/Macrophages eQTL
Monocytes/Macrophages sQTL
Microglia caQTL
Microglia eQTL
Microglia sQTL
Astrocytes eQTL
Excitatory Neurons eQTL
Inhibitory Neurons eQTL
Oligodendrocytes eQTL
OPCs eQTL
Endothelial Cells eQTL
COLOC PP4
0 0.4 0.7 1
Brain e/p/s/m/ha/caQTL
CSF pQTL
Blood/Plasma e/p/sQTL
Other Tissue e/sQTL
Cell-type Specific e/s/caQTL

**Supplement Figure S6. Gene prioritization at Tier-1 prioritized Loci.**

Summary of genetic colocalization (COLOC) analyses to prioritize potential causal genes at Tier-1 pleiotropic loci. Columns reporting findings for different QTL types (xQTL) across different tissues and cell-types. Top supported genes are listed per locus. For each category, the cell reports the best PP4 among the datasets collapsed within that category (e.g., “Other Tissue” corresponds to the best result across GTEx v8 tissues). Only COLOC findings with PP4>0.7 are displayed and contributed to gene prioritization. This figure contains all prioritized genes and features expanding on Figure.4 which only reports top prioritized genes.

| Rank | Motif | P-value | log P-pvalue | % of Targets | % of Background | STD(Bg STD) | Best Match/Details | Motif File |
|---|---|---|---|---|---|---|---|---|
| 1 | CGGCAGAGGTTT | 1e-12 | -2.917e+01 | 7.79% | 0.02% | 436.2bp (262.8bp) | GFI1B_HUMAN.H11MO.0.A(0.630) More Information \| Similar Motifs Found | motif file (matrix) |
| 2 * | TCGCGACGCC | 1e-11 | -2.622e+01 | 14.29% | 0.63% | 263.3bp (379.5bp) | KLF14_HUMAN.H11MO.0.D(0.605) More Information \| Similar Motifs Found | motif file (matrix) |
| 3 * | GGCCTCTTGCTT | 1e-11 | -2.601e+01 | 10.39% | 0.18% | 470.9bp (450.1bp) | MAX_HUMAN.H11MO.0.A(0.621) More Information \| Similar Motifs Found | motif file (matrix) |
| 4 * | CCGCCAGAGAGA | 1e-9 | -2.260e+01 | 10.39% | 0.29% | 258.6bp (339.9bp) | UBIP1_HUMAN.H11MO.0.D(0.624) More Information \| Similar Motifs Found | motif file (matrix) |
| 5 * | GCGACATGGC | 1e-9 | -2.206e+01 | 16.88% | 1.54% | 318.8bp (334.0bp) | CR3L1_HUMAN.H11MO.0.D(0.653) More Information \| Similar Motifs Found | motif file (matrix) |
| 6 * | TGTCCCTTTA | 1e-9 | -2.197e+01 | 22.08% | 3.15% | 357.5bp (397.1bp) | ANDR_HUMAN.H11MO.2.A(0.707) More Information \| Similar Motifs Found | motif file (matrix) |
| 7 * | GTAGTTAGTCGG | 1e-9 | -2.183e+01 | 7.79% | 0.09% | 207.3bp (290.2bp) | GLI2_HUMAN.H11MO.0.D(0.514) More Information \| Similar Motifs Found | motif file (matrix) |
| 8 * | CTGACGCCTT | 1e-9 | -2.132e+01 | 19.48% | 2.41% | 293.7bp (421.2bp) | ATF1_HUMAN.H11MO.0.B(0.647) More Information \| Similar Motifs Found | motif file (matrix) |
| 9 * | AGGCCGAGGTCC | 1e-9 | -2.110e+01 | 23.38% | 3.83% | 358.3bp (449.3bp) | ZFX_HUMAN.H11MO.1.A(0.683) More Information \| Similar Motifs Found | motif file (matrix) |
| 10 * | GAAGCTCTCTTT | 1e-8 | -2.066e+01 | 9.09% | 0.23% | 409.3bp (381.1bp) | NKX22_HUMAN.H11MO.0.D(0.617) More Information \| Similar Motifs Found | motif file (matrix) |
| 11 * | CCCTCAAACTTG | 1e-8 | -2.044e+01 | 10.39% | 0.39% | 332.6bp (372.0bp) | NKX21_HUMAN.H11MO.0.A(0.601) More Information \| Similar Motifs Found | motif file (matrix) |
| 12 * | GAGTTTGAAATA | 1e-8 | -2.038e+01 | 9.09% | 0.24% | 241.5bp (417.2bp) | HMGA1_HUMAN.H11MO.0.D(0.574) More Information \| Similar Motifs Found | motif file (matrix) |
| 13 * | TGTCGCGGAC | 1e-8 | -2.023e+01 | 16.88% | 1.80% | 261.2bp (423.4bp) | HINFP_HUMAN.H11MO.0.C(0.600) More Information \| Similar Motifs Found | motif file (matrix) |
| 14 * | ATGCGAGGACCC | 1e-8 | -1.961e+01 | 6.49% | 0.06% | 89.0bp (324.9bp) | ZBT7A_HUMAN.H11MO.0.A(0.585) More Information \| Similar Motifs Found | motif file (matrix) |
| 15 * | GGAGGGAGGGGA | 1e-8 | -1.878e+01 | 42.86% | 15.26% | 349.1bp (424.9bp) | PATZ1_HUMAN.H11MO.1.C(0.841) More Information \| Similar Motifs Found | motif file (matrix) |
| 16 * | GCAGTGGCTGAC | 1e-8 | -1.869e+01 | 7.79% | 0.16% | 257.6bp (441.1bp) | CEBPZ_HUMAN.H11MO.0.D(0.612) More Information \| Similar Motifs Found | motif file (matrix) |
| 17 * | GTCTTTTTTTGC | 1e-8 | -1.864e+01 | 6.49% | 0.07% | 377.9bp (371.8bp) | FUBP1_HUMAN.H11MO.0.D(0.748) More Information \| Similar Motifs Found | motif file (matrix) |
| 18 * | CGGGAGTTGT | 1e-8 | -1.858e+01 | 19.48% | 2.97% | 328.8bp (385.7bp) | PATZ1_HUMAN.H11MO.1.C(0.632) More Information \| Similar Motifs Found | motif file (matrix) |
| 19 * | ATTTGCAAAATC | 1e-7 | -1.759e+01 | 6.49% | 0.09% | 357.4bp (341.6bp) | PO3F4_HUMAN.H11MO.0.D(0.737) More Information \| Similar Motifs Found | motif file (matrix) |
| 20 * | GCTCTTAAAGGA | 1e-7 | -1.741e+01 | 11.69% | 0.85% | 375.7bp (361.0bp) | ZN667_HUMAN.H11MO.0.C(0.644) More Information \| Similar Motifs Found | motif file (matrix) |
| 21 * | AACCCCCCGTCT | 1e-7 | -1.673e+01 | 3.90% | 0.00% | 207.0bp (0.0bp) | PITX1_HUMAN.H11MO.0.D(0.621) More Information \| Similar Motifs Found | motif file (matrix) |
| 22 * | TCGTGGGACTGC | 1e-7 | -1.673e+01 | 3.90% | 0.00% | 163.8bp (0.0bp) | ARNT2_HUMAN.H11MO.0.D(0.602) More Information \| Similar Motifs Found | motif file (matrix) |
| 23 * | GTTGGTCTAGCA | 1e-7 | -1.668e+01 | 9.09% | 0.42% | 383.8bp (411.1bp) | HSF1_HUMAN.H11MO.1.A(0.559) More Information \| Similar Motifs Found | motif file (matrix) |
| 24 * | GGGGGATCCA | 1e-6 | -1.606e+01 | 15.58% | 2.14% | 230.1bp (436.8bp) | GLIS3_HUMAN.H11MO.0.D(0.651) More Information \| Similar Motifs Found | motif file (matrix) |
| 25 * | CCTGCGGGGAAG | 1e-6 | -1.566e+01 | 23.38% | 5.48% | 344.3bp (449.1bp) | AP2B_HUMAN.H11MO.0.B(0.672) More Information \| Similar Motifs Found | motif file (matrix) |
| 26 * | TCCGGGTGAG | 1e-6 | -1.552e+01 | 20.78% | 4.33% | 307.1bp (452.1bp) | MECP2_HUMAN.H11MO.0.C(0.680) More Information \| Similar Motifs Found | motif file (matrix) |
| 27 * | GTAGTAGATA | 1e-6 | -1.551e+01 | 6.49% | 0.15% | 274.5bp (237.1bp) | GATA2_HUMAN.H11MO.0.A(0.616) More Information \| Similar Motifs Found | motif file (matrix) |
| 28 * | TCATCATGAG | 1e-6 | -1.524e+01 | 9.09% | 0.53% | 282.7bp (381.4bp) | PO6F2_HUMAN.H11MO.0.D(0.608) More Information \| Similar Motifs Found | motif file (matrix) |
| 29 * | CGAGCAGTTC | 1e-6 | -1.519e+01 | 15.58% | 2.33% | 379.2bp (431.5bp) | ASCL2_HUMAN.H11MO.0.D(0.604) More Information \| Similar Motifs Found | motif file (matrix) |
| 30 * | AGCAACTTGGTG | 1e-6 | -1.487e+01 | 5.19% | 0.06% | 280.0bp (436.6bp) | SCRT1_HUMAN.H11MO.0.D(0.574) More Information \| Similar Motifs Found | motif file (matrix) |
| 31 * | CCAAGGTCAC | 1e-6 | -1.462e+01 | 12.99% | 1.57% | 313.7bp (418.5bp) | ERR1_HUMAN.H11MO.0.A(0.849) More Information \| Similar Motifs Found | motif file (matrix) |
| 32 * | GAGAGCGATG | 1e-6 | -1.449e+01 | 9.09% | 0.59% | 391.9bp (465.3bp) | NKX25_HUMAN.H11MO.0.B(0.586) More Information \| Similar Motifs Found | motif file (matrix) |
| 33 * | AAGGGGCCTG | 1e-6 | -1.439e+01 | 28.57% | 8.83% | 418.7bp (440.3bp) | ZFX_HUMAN.H11MO.0.A(0.776) More Information \| Similar Motifs Found | motif file (matrix) |
| 34 * | GGACCTCAGGTG | 1e-5 | -1.380e+01 | 10.39% | 0.97% | 400.9bp (441.7bp) | SNAI1_HUMAN.H11MO.0.C(0.751) More Information \| Similar Motifs Found | motif file (matrix) |
| 35 * | ACTTCCTCTT | 1e-5 | -1.287e+01 | 31.17% | 11.30% | 347.8bp (423.6bp) | SPI1_HUMAN.H11MO.0.A(0.797) More Information \| Similar Motifs Found | motif file (matrix) |
| 36 * | ATTACACGGG | 1e-5 | -1.267e+01 | 11.69% | 1.53% | 240.3bp (424.3bp) | FOXH1_HUMAN.H11MO.0.A(0.611) More Information \| Similar Motifs Found | motif file (matrix) |
| 37 * | GACTACAA | 1e-4 | -1.144e+01 | 12.99% | 2.27% | 382.8bp (335.0bp) | PEBB_HUMAN.H11MO.0.C(0.666) More Information \| Similar Motifs Found | motif file (matrix) |
| 38 * | TAGCATTGTCTA | 1e-4 | -1.114e+01 | 2.60% | 0.00% | 161.7bp (0.0bp) | SOX21_HUMAN.H11MO.0.D(0.558) More Information \| Similar Motifs Found | motif file (matrix) |
| 39 * | TCCTAAATCT | 1e-4 | -1.106e+01 | 6.49% | 0.39% | 323.1bp (392.2bp) | HXA9_HUMAN.H11MO.0.B(0.651) More Information \| Similar Motifs Found | motif file (matrix) |
| 40 * | CTGATGCAGGGA | 1e-3 | -7.825e+00 | 2.60% | 0.03% | 156.8bp (588.2bp) | ATF4_HUMAN.H11MO.0.A(0.621) More Information \| Similar Motifs Found | motif file (matrix) |
| 41 * | CAGTGATGGT | 1e-2 | -6.856e+00 | 10.39% | 2.65% | 259.4bp (432.9bp) | GFI1_HUMAN.H11MO.0.C(0.608) More Information \| Similar Motifs Found | motif file (matrix) |

**Supplement Figure S7. De novo transcription-factor motif enrichment (HOMER) for xQTL COLOC prioritized genes from all female-biased Tier-1 prioritized loci.**

Shown are the top non-redundant de novo motifs ranked by significance. Columns report Motif, P-value, log P-value, % of Targets, % of Background, STD (Bg STD), and Best Match/Details (HOMER/HOCOMOCO IDs with links to motif matrices). ANDR (androgen receptor/NR3C4) is among the top-ranked motifs (rank=6).

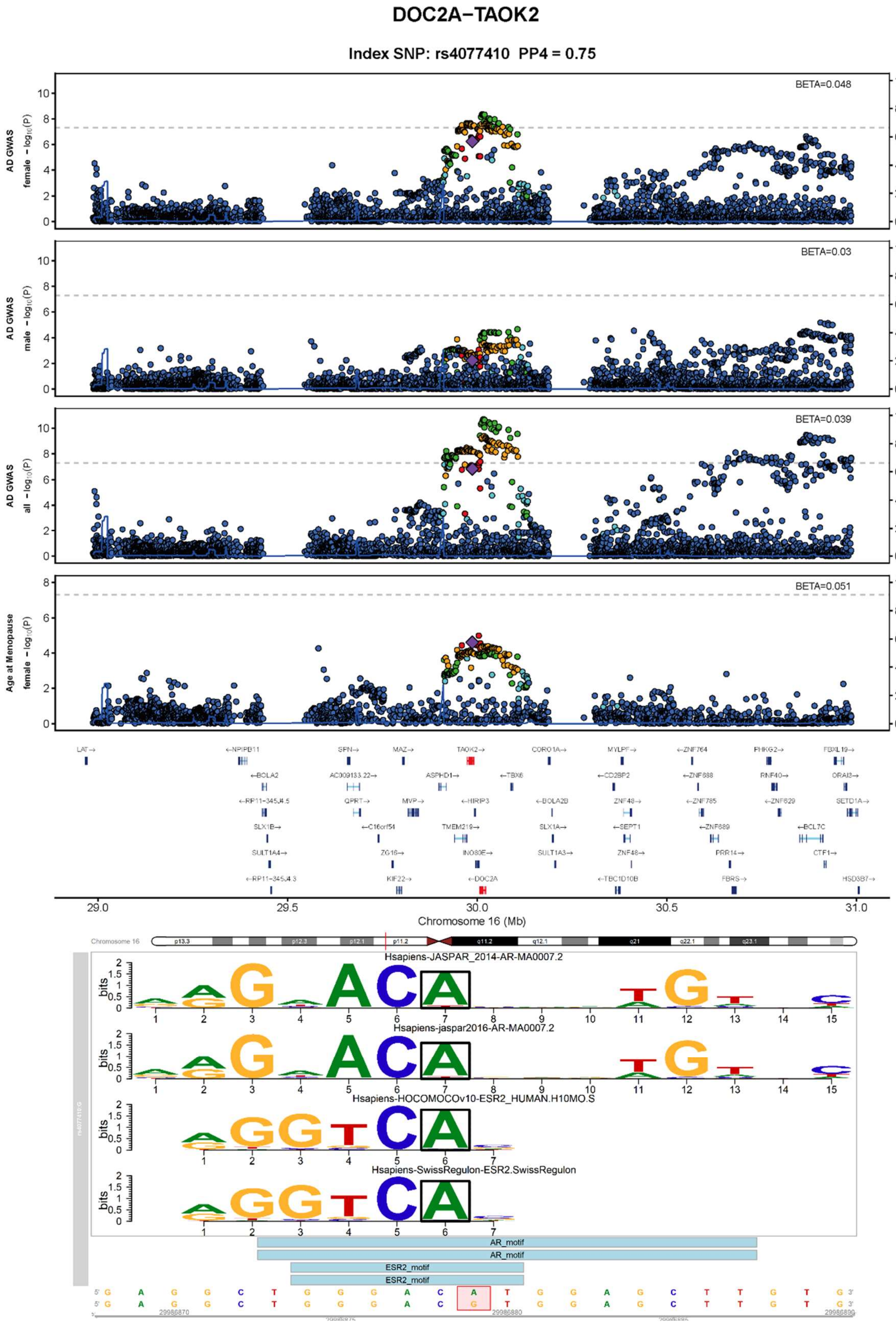

DOC2A−TAOK2
Index SNP: rs4077410 PP4 = 0.75
AD GWAS female −log10(P)
BETA=0.048
AD GWAS male −log10(P)
BETA=0.03
AD GWAS all −log10(P)
BETA=0.039
Age at Menopause female −log10(P)
BETA=0.051
Recombination rate (%)
LAT→
←NPIPB11
SPN→
MAZ→
TAOK2→
CORO1A→
MYLPF→
←ZNF764
PHKG2→
FBXL19→
←BOLA2
AC009133.22→
ASPHD1→
←TBX6
←CD2BP2
←ZNF688
RNF40→
ORAI3→
QPRT→
MVP→
←HIRIP3
←BOLA2B
ZNF48→
←ZNF785
←ZNF629
SETD1A→
SLX1B→
←C16orf54
TMEM219→
SLX1A→
←SEPT1
←ZNF689
←BCL7C
SULT1A4→
ZG16→
INO80E→
SULT1A3→
ZNF48→
PRR14→
CTF1→
KIF22→
←DOC2A
←TBC1D10B
FBRS→
HSD3B7→
29.0
29.5
30.0
30.5
31.0
Chromosome 16 (Mb)
Chromosome 16
Hsapiens-JASPAR_2014-AR-MA0007.2
Hsapiens-jaspar2016-AR-MA0007.2
Hsapiens-HOCOMOCOv10-ESR2_HUMAN.H10MO.S
Hsapiens-SwissRegulon-ESR2.SwissRegulon
bits
AR_motif
AR_motif
ESR2_motif
ESR2_motif
5' G A G G C T G G G A C A T G G A G C T T G T G 3'
5' G A G G C T G G G A C G T G G A G C T T G T G 3'
29986870
29986880
29986890

#### WDR81

##### Index SNP: rs62090040 PP4 = 0.95

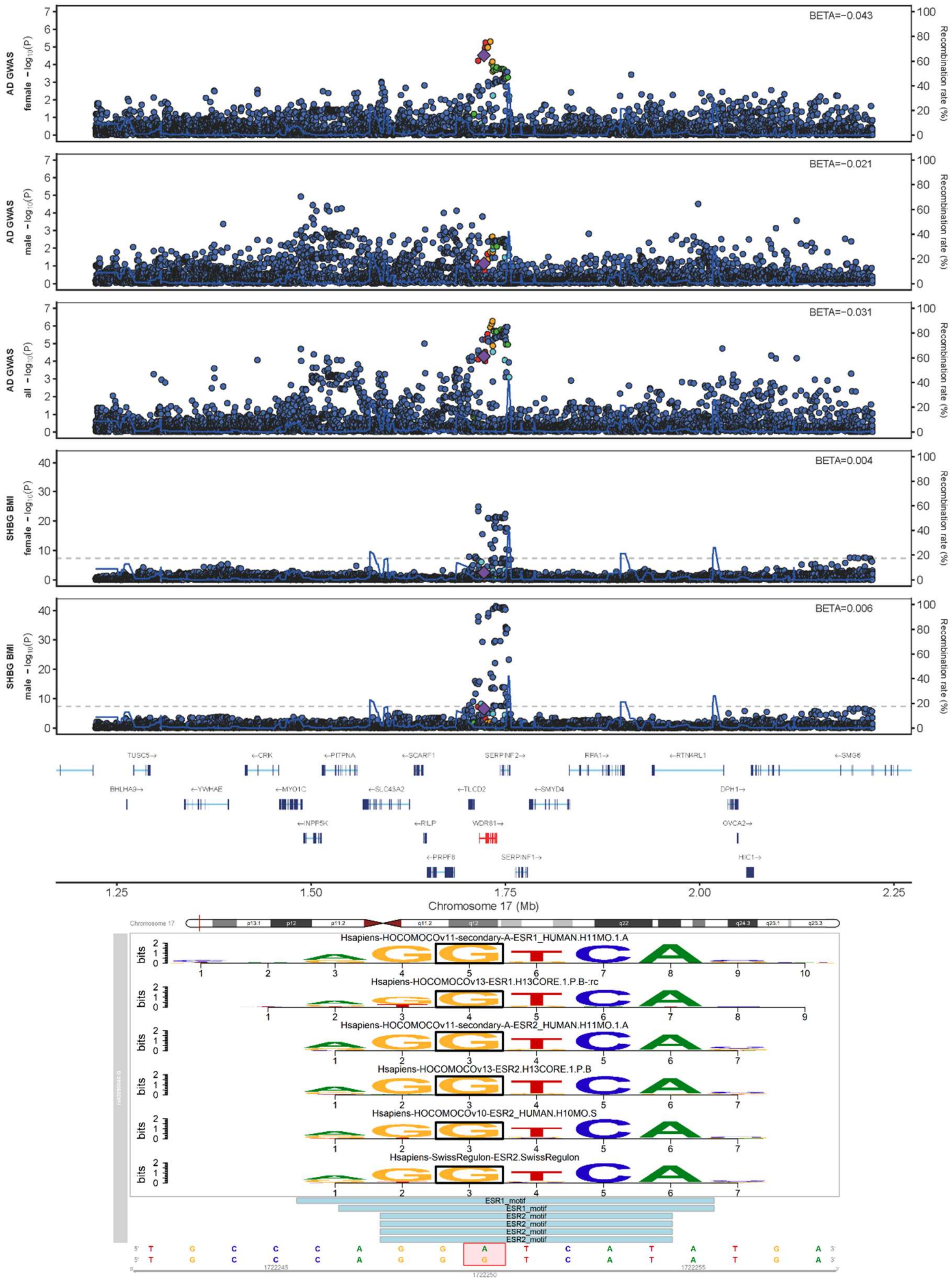

**Supplement Figure S8. Locus zoom and motif disruption plots for the two Tier-1 loci with significant motifbreakR support.**

A) DOC2A/TAOK2 locus. B) WDR81 locus. Top panels show locus patterns in AD and the corresponding sex hormone-related trait GWAS, while bottom panels show overlapping AR and ESR1/ESR2 motifs predicted by JASPAR, HOCOMOCO, and SwissRegulon. The highlighted variants are predicted to alter motif match scores, providing direct support for potential sex hormone regulatory effects at these loci.

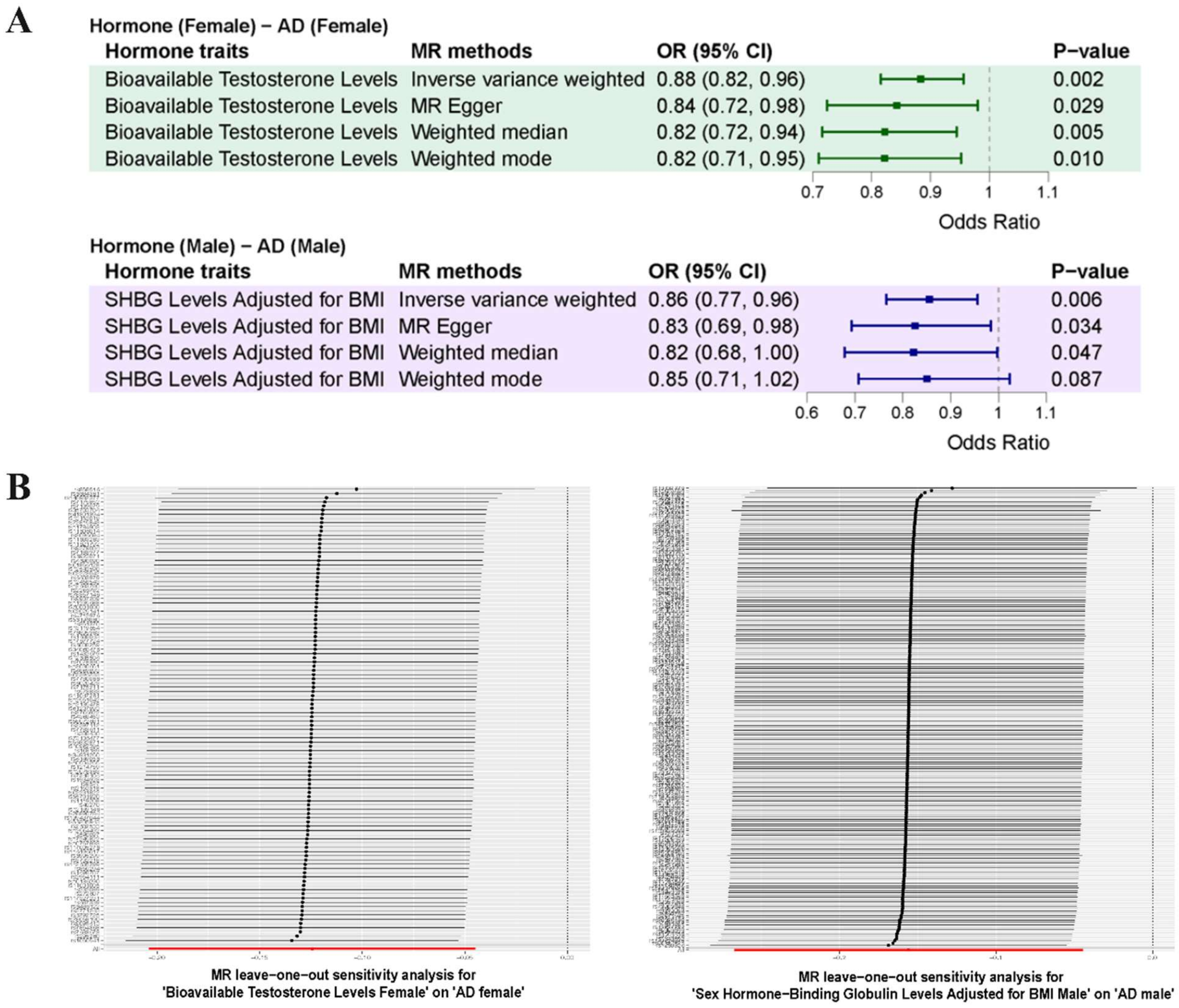

**Supplement Figure S9. MR sensitivity analyses for the two FDR-significant sex hormone and AD traits pairs.**

A) Forest plots for four MR estimators—inverse-variance weighted (IVW), MR-Egger, weighted median, and weighted mode—applied to Bioavailable testosterone level (female) → AD (female) and SHBG level adjusted for BMI (male) → AD (male). Points show odds ratios (OR) with 95% CIs; the dashed line marks OR = 1; P-values are reported. B) Leave-one-out analyses for the instrument sets matching (A). Each row shows the IVW estimate after removing one SNP; the solid vertical line indicates the full IVW estimate

### Acknowledgment

Data for this study were prepared, archived, and distributed by the National Institute on Aging Alzheimer's Disease Data Storage Site (NIAGADS) at the University of Pennsylvania (U24-AG041689), funded by the National Institute on Aging. The contents of this article do not represent the views of the National Institutes of Health, the U.S. Department of Veterans Affairs, or the United States Government.

#### Acknowledgments for the use of ADSP data

The Alzheimer's Disease Sequencing Project (ADSP) is comprised of two Alzheimer's Disease (AD) genetics consortia and three National Human Genome Research Institute (NHGRI) funded Large Scale Sequencing and Analysis Centers (LSAC). The two AD genetics consortia are the Alzheimer's Disease Genetics Consortium (ADGC) funded by NIA (U01 AG032984), and the Cohorts for Heart and Aging Research in Genomic Epidemiology (CHARGE) funded by NIA (R01 AG033193), the National Heart, Lung, and Blood Institute (NHLBI), other National Institute of Health (NIH) institutes and other foreign governmental and non-governmental organizations. The Discovery Phase analysis of sequence data is supported through UF1AG047133 (to Drs. Schellenberg, Farrer, Pericak-Vance, Mayeux, and Haines); U01AG049505 to Dr. Seshadri; U01AG049506 to Dr. Boerwinkle; U01AG049507 to Dr. Wijsman; and U01AG049508 to Dr. Goate and the Discovery Extension Phase analysis is supported through U01AG052411 to Dr. Goate, U01AG052410 to Dr. Pericak-Vance and U01 AG052409 to Drs. Seshadri and Fornage.

Sequencing for the Follow Up Study (FUS) is supported through U01AG057659 (to Drs. PericakVance, Mayeux, and Vardarajan) and U01AG062943 (to Drs. Pericak-Vance and Mayeux). Data generation and harmonization in the Follow-up Phase is supported by U54AG052427 (to Drs. Schellenberg and Wang). The FUS Phase analysis of sequence data is supported through U01AG058589 (to Drs. Destefano, Boerwinkle, De Jager, Fornage, Seshadri, and Wijsman), U01AG058654 (to Drs. Haines, Bush, Farrer, Martin, and Pericak-Vance), U01AG058635 (to Dr. Goate), RF1AG058066 (to Drs. Haines, Pericak-Vance, and Scott), RF1AG057519 (to Drs. Farrer and Jun), R01AG048927 (to Dr. Farrer), and RF1AG054074 (to Drs. Pericak-Vance and Beecham).

The ADGC cohorts include: Adult Changes in Thought (ACT) (UO1 AG006781, UO1 HG004610, UO1 HG006375, U01 HG008657), the Alzheimer's Disease Centers (ADC) ( P30 AG019610, P30 AG013846, P50 AG008702, P50 AG025688, P50 AG047266, P30 AG010133, P50 AG005146, P50 AG005134, P50 AG016574, P50 AG005138, P30 AG008051, P30 AG013854, P30 AG008017, P30 AG010161, P50 AG047366, P30 AG010129, P50 AG016573, P50 AG016570, P50 AG005131, P50 AG023501, P30 AG035982, P30 AG028383, P30 AG010124, P50 AG005133, P50 AG005142, P30 AG012300, P50 AG005136, P50 AG033514, P50 AG005681, and P50 AG047270), the Chicago Health and Aging Project (CHAP) (R01 AG11101, RC4

AG039085, K23 AG030944), Indianapolis Ibadan (R01 AG009956, P30 AG010133), the Memory and Aging Project (MAP) ( R01 AG17917), Mayo Clinic (MAYO) (R01 AG032990, U01 AG046139, R01 NS080820, RF1 AG051504, P50 AG016574), Mayo Parkinson's Disease controls (NS039764, NS071674, 5RC2HG005605), University of Miami (R01 AG027944, R01 AG028786, R01 AG019085, IIRG09133827, A2011048), the Multi-Institutional Research in Alzheimer's Genetic Epidemiology Study (MIRAGE) (R01 AG09029, R01 AG025259), the National Cell Repository for Alzheimer's Disease (NCRAD) (U24 AG21886), the National Institute on Aging Late Onset Alzheimer's Disease Family Study (NIA- LOAD) (R01 AG041797), the Religious Orders Study (ROS) (P30 AG10161, R01 AG15819), the Texas Alzheimer's Research and Care Consortium (TARCC) (funded by the Darrell K Royal Texas Alzheimer's Initiative), Vanderbilt University/Case Western Reserve University (VAN/CWRU) (R01 AG019757, R01 AG021547, R01 AG027944, R01 AG028786, P01 NS026630, and Alzheimer's Association), the Washington Heights-Inwood Columbia Aging Project (WHICAP) (RF1 AG054023), the University of Washington Families (VA Research Merit Grant, NIA: P50AG005136, R01AG041797, NINDS: R01NS069719), the Columbia University HispanicEstudio Familiar de Influencia Genetica de Alzheimer (EFIGA) (RF1 AG015473), the University of Toronto (UT) (funded by Wellcome Trust, Medical Research Council, Canadian Institutes of Health Research), and Genetic Differences (GD) (R01 AG007584). The CHARGE cohorts are supported in part by National Heart, Lung, and Blood Institute (NHLBI) infrastructure grant HL105756 (Psaty), RC2HL102419 (Boerwinkle) and the neurology working group is supported by the National Institute on Aging (NIA) R01 grant AG033193.

The CHARGE cohorts participating in the ADSP include the following: Austrian Stroke Prevention Study (ASPS), ASPS-Family study, and the Prospective Dementia Registry-Austria (ASPS/PRODEM-Aus), the Atherosclerosis Risk in Communities (ARIC) Study, the Cardiovascular Health Study (CHS), the Erasmus Rucphen Family Study (ERF), the Framingham Heart Study (FHS), and the Rotterdam Study (RS). ASPS is funded by the Austrian Science Fond (FWF) grant number P20545-P05 and P13180 and the Medical University of Graz. The ASPS-Fam is funded by the Austrian Science Fund (FWF) project I904),the EU Joint Programme - Neurodegenerative Disease Research (JPND) in frame of the BRIDGET project (Austria, Ministry of Science) and the Medical University of Graz and the Steiermärkische Krankenanstalten Gesellschaft. PRODEM-Austria is supported by the Austrian Research Promotion agency (FFG) (Project No. 827462) and by the Austrian National Bank (Anniversary Fund, project 15435. ARIC research is carried out as a collaborative study supported by NHLBI contracts (HHSN268201100005C, HHSN268201100006C, HHSN268201100007C, HHSN268201100008C, HHSN268201100009C, HHSN268201100010C, HHSN268201100011C, and HHSN268201100012C). Neurocognitive data in ARIC is collected by U01 2U01HL096812, 2U01HL096814, 2U01HL096899, 2U01HL096902, 2U01HL096917 from the NIH (NHLBI, NINDS, NIA and NIDCD), and with previous brain MRI examinations funded by R01-HL70825 from the NHLBI. CHS research was supported by contracts HHSN268201200036C, HHSN268200800007C, N01HC55222, N01HC85079, N01HC85080, N01HC85081, N01HC85082, N01HC85083, N01HC85086, and grants U01HL080295 and U01HL130114 from the NHLBI with additional contribution from the National Institute of Neurological Disorders and Stroke (NINDS). Additional support was provided by R01AG023629, R01AG15928, and R01AG20098 from the NIA. FHS research is supported by NHLBI contracts N01-HC-25195 and HHSN268201500001I. This study was also supported by additional grants from the NIA (R01s AG054076, AG049607 and AG033040 and NINDS (R01 NS017950). The ERF study as a part of EUROSPAN (European Special Populations Research Network) was supported by European Commission FP6 STRP grant number 018947 (LSHG-CT-2006-01947) and also received funding from the European Community's Seventh Framework Programme (FP7/2007-2013)/grant agreement HEALTH-F4- 2007-201413 by the European Commission under the programme "Quality of Life and Management of the Living Resources" of 5th Framework Programme (no. QLG2-CT-2002- 01254). High-throughput analysis of the ERF data was supported by a joint grant from the Netherlands Organization for Scientific Research and the Russian Foundation for Basic Research (NWO-RFBR 047.017.043). The Rotterdam Study is funded by Erasmus Medical Center and Erasmus University, Rotterdam, the Netherlands Organization for Health Research and Development (ZonMw), the Research Institute for Diseases in the Elderly (RIDE), the Ministry of Education, Culture and Science, the Ministry for Health, Welfare and Sports, the European Commission (DG XII), and the municipality of Rotterdam. Genetic data sets are also supported by the Netherlands Organization of Scientific Research NWO Investments (175.010.2005.011, 911-03-012), the Genetic Laboratory of the Department of Internal Medicine, Erasmus MC, the Research Institute for Diseases in the Elderly (014-93-015; RIDE2), and the Netherlands Genomics Initiative (NGI)/Netherlands Organization for Scientific Research (NWO) Netherlands Consortium for Healthy Aging (NCHA), project 050-060-810. All studies are grateful to their participants, faculty and staff. The content of these manuscripts is solely the responsibility of the authors and does not necessarily represent the official views of the National Institutes of Health or the U.S. Department of Health and Human Services.

The FUS cohorts include: the Alzheimer's Disease Centers (ADC) ( P30 AG019610, P30 AG013846, P50 AG008702, P50 AG025688, P50 AG047266, P30 AG010133, P50 AG005146, P50 AG005134, P50 AG016574, P50 AG005138, P30 AG008051, P30 AG013854, P30 AG008017, P30 AG010161, P50 AG047366, P30 AG010129, P50 AG016573, P50 AG016570, P50 AG005131, P50 AG023501, P30 AG035982, P30 AG028383, P30 AG010124, P50 AG005133, P50 AG005142, P30 AG012300, P50 AG005136, P50 AG033514, P50 AG005681, and P50 AG047270), Alzheimer's Disease Neuroimaging Initiative (ADNI) (U19AG024904), Amish Protective Variant Study (RF1AG058066), Cache County Study (R01AG11380, R01AG031272, R01AG21136, RF1AG054052), Case Western Reserve University Brain Bank (CWRUBB) (P50AG008012), Case Western Reserve University Rapid Decline (CWRURD) (RF1AG058267, NU38CK000480), CubanAmerican Alzheimer's Disease Initiative (CuAADI) (3U01AG052410), Estudio Familiar de Influencia Genetica en Alzheimer (EFIGA) (5R37AG015473, RF1AG015473, R56AG051876), Genetic and Environmental Risk Factors for Alzheimer Disease Among African Americans Study (GenerAAtions) (2R01AG09029, R01AG025259, 2R01AG048927), Gwangju Alzheimer and Related Dementias Study (GARD) (U01AG062602), Hussman Institute for Human Genomics Brain Bank (HIHGBB) (R01AG027944, Alzheimer's Association "Identification of Rare Variants in Alzheimer Disease"), Ibadan Study of Aging (IBADAN) (5R01AG009956), Mexican Health and Aging Study (MHAS) (R01AG018016), Multi-Institutional Research in Alzheimer's Genetic Epidemiology (MIRAGE) (2R01AG09029, R01AG025259, 2R01AG048927), Northern Manhattan Study (NOMAS) (R01NS29993), Peru Alzheimer's Disease Initiative (PeADI) (RF1AG054074), Puerto Rican 1066 (PR1066) (Wellcome Trust (GR066133/GR080002), European Research Council (340755)), Puerto Rican Alzheimer Disease Initiative (PRADI) (RF1AG054074), Reasons for Geographic and Racial Differences in Stroke (REGARDS) (U01NS041588), Research in African American Alzheimer Disease Initiative (REAAADI) (U01AG052410), Rush Alzheimer's Disease Center (ROSMAP) (P30AG10161, R01AG15819, R01AG17919), University of Miami Brain Endowment Bank (MBB), and

University of Miami/Case Western/North Carolina A&T African American (UM/CASE/NCAT) (U01AG052410, R01AG028786).

The four LSACs are: the Human Genome Sequencing Center at the Baylor College of Medicine (U54 HG003273), the Broad Institute Genome Center (U54HG003067), The American Genome Center at the Uniformed Services University of the Health Sciences (U01AG057659), and the Washington University Genome Institute (U54HG003079).

Biological samples and associated phenotypic data used in primary data analyses were stored at Study Investigators institutions, and at the National Cell Repository for Alzheimer's Disease (NCRAD, U24AG021886) at Indiana University funded by NIA. Associated Phenotypic Data used in primary and secondary data analyses were provided by Study Investigators, the NIA funded Alzheimer's Disease Centers (ADCs), and the National Alzheimer's Coordinating Center (NACC, U01AG016976) and the National Institute on Aging Genetics of Alzheimer's Disease Data Storage Site (NIAGADS, U24AG041689) at the University of Pennsylvania, funded by NIA This research was supported in part by the Intramural Research Program of the National Institutes of health, National Library of Medicine. Contributors to the Genetic Analysis Data included Study Investigators on projects that were individually funded by NIA, and other NIH institutes, and by private U.S. organizations, or foreign governmental or nongovernmental organizations.

An up to date acknowledgment statement can be found on the ADSP site: https://www.niagads.org/adsp/content/acknowledgement-statement.

Data collection and sharing for this project was funded by the Alzheimer's Disease Neuroimaging Initiative (ADNI) (National Institutes of Health Grant U01 AG024904) and DOD ADNI (Department of Defense award number W81XWH-12-2-0012). ADNI is funded by the National Institute on Aging, the National Institute of Biomedical Imaging and Bioengineering, and through generous contributions from the following: AbbVie, Alzheimer's Association; Alzheimer's Drug Discovery Foundation; Araclon Biotech; BioClinica, Inc.; Biogen; Bristol-Myers Squibb Company; CereSpir, Inc.; Cogstate; Eisai Inc.; Elan Pharmaceuticals, Inc.; Eli Lilly and Company; EuroImmun; F. Hoffmann-La Roche Ltd and its affiliated company Genentech, Inc.; Fujirebio; GE Healthcare; IXICO Ltd.; Janssen Alzheimer Immunotherapy Research & Development, LLC.; Johnson & Johnson Pharmaceutical Research & Development LLC.; Lumosity; Lundbeck; Merck & Co., Inc.; Meso Scale Diagnostics, LLC.; NeuroRx Research; Neurotrack Technologies; Novartis Pharmaceuticals Corporation; Pfizer Inc.; Piramal Imaging; Servier; Takeda Pharmaceutical Company; and Transition Therapeutics. The Canadian Institutes of Health Research is providing funds to support ADNI clinical sites in Canada. Private sector contributions are facilitated by the Foundation for the National Institutes of Health (www.fnih.org). The grantee organization is the Northern California Institute for Research and Education, and the study is coordinated by the Alzheimer's Therapeutic Research Institute at the University of Southern California. ADNI data are disseminated by the Laboratory for Neuro Imaging at the University of Southern California.

Additional information to include in an acknowledgment statement can be found on the LONI site: https://adni.loni.usc.edu/wp-content/uploads/how_to_apply/ADNI_Data_Use_Agreement.pdf.

The Alzheimer's Disease Genetics Consortium (ADGC) supported sample preparation, whole exome sequencing and data processing through NIA grant U01AG032984. Sequencing data generation and harmonization is supported by the Genome Center for Alzheimer's Disease, U54AG052427, and data sharing is supported by NIAGADS, U24AG041689. Samples from the National Centralized Repository for Alzheimer's Disease and Related Dementias (NCRAD), which receives government support under a cooperative agreement grant (U24 AG021886) awarded by the National Institute on Aging (NIA), were used in this study. We thank contributors who collected samples used in this study, as well as patients and their families, whose help and participation made this work possible. NIH grants supported enrollment and data collection for the individual studies including: GenerAAtions R01AG20688 (PI M. Daniele Fallin, PhD); Miami/Duke R01 AG027944, R01 AG028786 (PI Margaret A. Pericak-Vance, PhD); NC A&T P20 MD000546, R01 AG28786-01A1 (PI Goldie S. Byrd, PhD); Case Western (PI Jonathan L. Haines, PhD); MIRAGE R01 AG009029 (PI Lindsay A. Farrer, PhD); ROS P30AG10161, R01AG15819, R01AG30146, TGen (PI David A. Bennett, MD); MAP R01AG17917, R01AG15819, TGen (PI David A. Bennett, MD). The NACC database is funded by NIA/NIH Grant U01 AG016976. NACC data are contributed by the NIA-funded ADCs: P30 AG019610 (PI Eric Reiman, MD), P30 AG013846 (PI Neil Kowall, MD), P30 AG062428-01 (PI James Leverenz, MD) P50 AG008702 (PI Scott Small, MD), P50 AG025688 (PI Allan Levey, MD, PhD), P50 AG047266 (PI Todd Golde, MD, PhD), P30 AG010133 (PI Andrew Saykin, PsyD), P50 AG005146 (PI Marilyn Albert, PhD), P30 AG062421-01 (PI Bradley Hyman, MD, PhD), P30 AG062422-01 (PI Ronald Petersen, MD, PhD), P50 AG005138 (PI Mary Sano, PhD), P30 AG008051 (PI Thomas Wisniewski, MD), P30 AG013854 (PI Robert Vassar, PhD), P30 AG008017 (PI Jeffrey Kaye, MD), P30 AG010161 (PI David Bennett, MD), P50 AG047366 (PI Victor Henderson, MD, MS), P30 AG010129 (PI Charles DeCarli, MD), P50 AG016573 (PI Frank LaFerla, PhD), P30 AG062429-01(PI James Brewer, MD, PhD), P50 AG023501 (PI Bruce Miller, MD), P30 AG035982 (PI Russell Swerdlow, MD), P30 AG028383 (PI Linda Van Eldik, PhD), P30 AG053760 (PI Henry Paulson, MD, PhD), P30 AG010124 (PI John Trojanowski, MD, PhD), P50 AG005133 (PI Oscar Lopez, MD), P50 AG005142 (PI Helena Chui, MD), P30 AG012300 (PI Roger Rosenberg, MD), P30 AG049638 (PI Suzanne Craft, PhD), P50 AG005136 (PI Thomas Grabowski, MD), P30 AG062715-01 (PI Sanjay Asthana, MD, FRCP), P50 AG005681 (PI John Morris, MD), P50 AG047270 (PI Stephen Strittmatter, MD, PhD).

This work was supported by grants from the National Institutes of Health (R01AG044546, P01AG003991, RF1AG053303, R01AG058501, U01AG058922, RF1AG058501 and R01AG057777). The recruitment and clinical characterization of research participants at Washington University were supported by NIH P50 AG05681, P01 AG03991, and P01 AG026276. This work was supported by access to equipment made possible by the Hope Center for Neurological Disorders, and the Departments of Neurology and Psychiatry at Washington University School of Medicine.

We thank the contributors who collected samples used in this study, as well as patients and their families, whose help and participation made this work possible. Members of the National Institute on Aging Late-Onset Alzheimer Disease/National Cell Repository for Alzheimer Disease (NIA-LOAD NCRAD) Family Study Group include the following: Richard Mayeux, MD, MSc; Martin Farlow, MD; Tatiana Foroud, PhD; Kelley Faber, MS; Bradley F. Boeve, MD; Neill R. Graff-Radford, MD; David A. Bennett, MD; Robert A. Sweet, MD; Roger Rosenberg, MD; Thomas D. Bird, MD; Carlos Cruchaga, PhD; and Jeremy M. Silverman, PhD.

This work was partially supported by grant funding from NIH R01 AG039700 and NIH P50 AG005136. Subjects and samples used here were originally collected with grant funding from NIH U24 AG026395, U24 AG021886, P50 AG008702, P01 AG007232, R37 AG015473, P30 AG028377, P50 AG05128, P50 AG16574,

P30 AG010133, P50 AG005681, P01 AG003991, U01MH046281, U01 MH046290 and U01 MH046373. The funders had no role in study design, analysis or preparation of the manuscript. The authors declare no competing interests.

This work was supported by the National Institutes of Health (R01 AG027944, R01 AG028786 to MAPV, R01 AG019085 to JLH, P20 MD000546); a joint grant from the Alzheimer's Association (SG-14-312644) and the Fidelity Biosciences Research Initiative to MAPV; the BrightFocus Foundation (A2011048 to MAPV). NIA-LOAD Family-Based Study supported the collection of samples used in this study through NIH grants U24 AG026395 and R01 AG041797 and the MIRAGE cohort was supported through the NIH grants R01 AG025259 and R01 AG048927. We thank contributors, including the Alzheimer's disease Centers who collected samples used in this study, as well as patients and their families, whose help and participation made this work possible. Study design: HNC, BWK, JLH, MAPV; Sample collection: MLC, JMV, RMC, LAF, JLH, MAPV; Whole exome sequencing and Sanger sequencing: SR, PLW; Sequencing data analysis: HNC, BWK, KLHN, SR, MAK, JRG, ERM, GWB, MAPV; Statistical analysis: BWK, KLHN, JMJ, MAPV; Preparation of manuscript: HNC, BWK. The authors jointly discussed the experimental results throughout the duration of the study. All authors read and approved the final manuscript.

Data collection and sharing for this project was supported by the Washington Heights-Inwood Columbia Aging Project (WHICAP, PO1AG07232, R01AG037212, RF1AG054023) funded by the National Institute on Aging (NIA) and by the National Center for Advancing Translational Sciences, National Institutes of Health, through Grant Number UL1TR001873. This manuscript has been reviewed by WHICAP investigators for scientific content and consistency of data interpretation with previous WHICAP Study publications. We acknowledge the WHICAP study participants and the WHICAP research and support staff for their contributions to this study.

This work was supported by grants from the National Institutes of Health (R01AG044546, P01AG003991, RF1AG053303, R01AG058501, U01AG058922, RF1AG058501 and R01AG057777). The recruitment and clinical characterization of research participants at Washington University were supported by NIH P50 AG05681, P01 AG03991, and P01 AG026276. This work was supported by access to equipment made possible by the Hope Center for Neurological Disorders, and the Departments of Neurology and Psychiatry at Washington University School of Medicine.

We thank the contributors who collected samples used in this study, as well as patients and their families, whose help and participation made this work possible. Members of the National Institute on Aging Late-Onset Alzheimer Disease/National Cell Repository for Alzheimer Disease (NIA-LOAD NCRAD) Family Study Group include the following: Richard Mayeux, MD, MSc; Martin Farlow, MD; Tatiana Foroud, PhD; Kelley Faber, MS; Bradley F. Boeve, MD; Neill R. Graff-Radford, MD; David A. Bennett, MD; Robert A. Sweet, MD; Roger Rosenberg, MD; Thomas D. Bird, MD; Carlos Cruchaga, PhD; and Jeremy M. Silverman, PhD.

This work was supported by grants from the National Institutes of Health (R01AG044546, P01AG003991, RF1AG053303, R01AG058501, U01AG058922, RF1AG058501 and R01AG057777). The recruitment and clinical characterization of research participants at Washington University were supported by NIH P50 AG05681, P01 AG03991, and P01 AG026276. This work was supported by access to equipment made possible by the Hope Center for Neurological Disorders, and the Departments of Neurology and Psychiatry at Washington University School of Medicine.

We thank the contributors who collected samples used in this study, as well as patients and their families, whose help and participation made this work possible. Members of the National Institute on Aging Late-Onset Alzheimer Disease/National Cell Repository for Alzheimer Disease (NIA-LOAD NCRAD) Family Study Group include the following: Richard Mayeux, MD, MSc; Martin Farlow, MD; Tatiana Foroud, PhD; Kelley Faber, MS; Bradley F. Boeve, MD; Neill R. Graff-Radford, MD; David A. Bennett, MD; Robert A. Sweet, MD; Roger Rosenberg, MD; Thomas D. Bird, MD; Carlos Cruchaga, PhD; and Jeremy M. Silverman, PhD.

Mayo RNAseq Study- Study data were provided by the following sources: The Mayo Clinic Alzheimer's Disease Genetic Studies, led by Dr. Nilufer Ertekin-Taner and Dr. Steven G. Younkin, Mayo Clinic, Jacksonville, FL using samples from the Mayo Clinic Study of Aging, the Mayo Clinic Alzheimer's Disease Research Center, and the Mayo Clinic Brain Bank. Data collection was supported through funding by NIA grants P50 AG016574, R01 AG032990, U01 AG046139, R01 AG018023, U01 AG006576, U01 AG006786, R01 AG025711, R01 AG017216, R01 AG003949, NINDS grant R01 NS080820, CurePSP Foundation, and support from Mayo Foundation. Study data includes samples collected through the Sun Health Research Institute Brain and Body Donation Program of Sun City, Arizona. The Brain and Body Donation Program is supported by the National Institute of Neurological Disorders and Stroke (U24 NS072026 National Brain and Tissue Resource for Parkinson's Disease and Related Disorders), the National Institute on Aging (P30 AG19610 Arizona Alzheimer's Disease Core Center), the Arizona Department of Health Services (contract 211002, Arizona Alzheimer's Research Center), the Arizona Biomedical Research Commission (contracts 4001, 0011, 05-901 and 1001 to the Arizona Parkinson's Disease Consortium) and the Michael J. Fox Foundation for Parkinson's Research

ROSMAP- We are grateful to the participants in the Religious Order Study, the Memory and Aging Project. This work is supported by the US National Institutes of Health [U01 AG046152, R01 AG043617, R01 AG042210, R01 AG036042, R01 AG036836, R01 AG032990, R01 AG18023, RC2 AG036547, P50 AG016574, U01 ES017155, KL2 RR024151, K25 AG041906-01, R01 AG30146, P30 AG10161, R01 AG17917, R01 AG15819, K08 AG034290, P30 AG10161 and R01 AG11101.

Mount Sinai Brain Bank (MSBB)- This work was supported by the grants R01AG046170, RF1AG054014, RF1AG057440 and R01AG057907 from the NIH/National Institute on Aging (NIA). R01AG046170 is a component of the AMP-AD Target Discovery and Preclinical Validation Project. Brain tissue collection and characterization was supported by NIH HHSN271201300031C.

This study was supported by the National Institute on Aging (NIA) grants AG030653, AG041718, AG064877 and P30-AG066468.

We would like to thank study participants, their families, and the sample collectors for their invaluable contributions. This research was supported in part by the National Institute on Aging grant U01AG049508 (PI Alison M. Goate). This research was supported in part by Genentech, Inc. (PI Alison M. Goate, Robert R. Graham).

The NACC database is funded by NIA/NIH Grant U01 AG016976. NACC data are contributed by these NIA-funded ADCs: P30 AG013846 (PI Neil Kowall, MD), P50 AG008702 (PI Scott Small, MD), P50 AG025688 (PI Allan Levey, MD, PhD), P30 AG010133 (PI Andrew Saykin, PsyD), P50 AG005146 (PI Marilyn Albert, PhD), P50 AG005134 (PI Bradley Hyman, MD, PhD), P50 AG016574 (PI Ronald Petersen, MD, PhD), P30 AG013854 (PI M. Marsel Mesulam, MD), P30 AG008017 (PI Jeffrey Kaye, MD), P30 AG010161 (PI David Bennett, MD), P30 AG010129 (PI Charles DeCarli, MD), P50 AG016573 (PI Frank LaFerla, PhD), P50 AG005131 (PI Douglas Galasko, MD), P30 AG028383 (PI Linda Van Eldik, PhD), P30 AG010124 (PI John Trojanowski, MD, PhD), P50 AG005142 (PI Helena Chui, MD), P30 AG012300 (PI Roger Rosenberg, MD), P50 AG005136 (PI Thomas Grabowski, MD), P50 AG005681 (PI John Morris, MD), P30 AG028377 (Kathleen Welsh-Bohmer, PhD), and P50 AG008671 (PI Henry Paulson, MD, PhD).

Samples from the National Cell Repository for Alzheimer's Disease (NCRAD), which receives government support under a cooperative agreement grant (U24 AG21886) awarded by the National Institute on Aging (NIA), were used in this study. We thank contributors who collected samples used in this study, as well as patients and their families, whose help and participation made this work possible.

The Alzheimer's Disease Genetics Consortium supported the collection of samples used in this study through National Institute on Aging (NIA) grants U01AG032984 and RC2AG036528.

We acknowledge the generous contributions of the Cache County Memory Study participants. Sequencing for this study was funded by RF1AG054052 (PI: John S.K. Kauwe)

**Acknowledgments for the use of GWAS data distributed by NIAGADS**

The NIA Genetics of Alzheimer's Disease Data Storage Site (NIAGADS) is supported by a collaborative agreement from the National Institute on Aging, U24AG041689.

NG00047: The NIA supported this work through grants U01-AG032984, RC2-AG036528, U01-AG016976 (Dr Kukull); U24 AG026395, U24 AG026390, R01AG037212, R37 AG015473 (Dr Mayeux); K23AG034550 (Dr Reitz); U24-AG021886 (Dr Foroud); R01AG009956, RC2 AG036650 (Dr Hall); UO1 AG06781, UO1 HG004610 (Dr Larson); R01 AG009029 (Dr Farrer); 5R01AG20688 (Dr Fallin); P50 AG005133, AG030653 (Dr Kamboh); R01 AG019085 (Dr Haines); R01 AG1101, R01 AG030146, RC2 AG036650 (Dr Evans); P30AG10161, R01AG15819, R01AG30146, R01AG17917, R01AG15819 (Dr Bennett); R01AG028786 (Dr Manly); R01AG22018, P30AG10161 (Dr Barnes); P50AG16574 (Dr Ertekin-Taner, Dr Graff-Radford), R01 AG032990 (Dr Ertekin-Taner), KL2 RR024151 (Dr Ertekin-Taner); R01 AG027944, R01 AG028786 (Dr Pericak-Vance); P20 MD000546, R01 AG28786-01A1 (Dr Byrd); AG005138 (Dr Buxbaum); P50 AG05681, P01 AG03991, P01 AG026276 (Dr Goate); and P30AG019610, P30AG13846, U01-AG10483, R01CA129769, R01MH080295, R01AG017173, R01AG025259, R01AG33193, P50AG008702, P30AG028377, AG05128, AG025688, P30AG10133, P50AG005146, P50AG005134, P01AG002219, P30AG08051, MO1RR00096, UL1RR029893, P30AG013854, P30AG008017, R01AG026916, R01AG019085, P50AG016582, UL1RR02777, R01AG031581, P30AG010129, P50AG016573, P50AG016575, P50AG016576, P50AG016577, P50AG016570, P50AG005131, P50AG023501, P50AG019724, P30AG028383, P50AG008671, P30AG010124, P50AG005142, P30AG012300, AG010491, AG027944, AG021547, AG019757,

P50AG005136 (Alzheimer Disease GeneticsConsortium [ADGC]). We thank Creighton Phelps, Stephen Synder, and Marilyn Miller from the NIA, who are ex-officio members of the ADGC. Support was also provided by the Alzheimer's Association (IIRG-08-89720 [Dr Farrer] and IIRG-05-14147 [Dr Pericak-Vance]), National Institute of Neurological Disorders and Stroke grant NS39764, National Institute of Mental Health grant MH60451, GlaxoSmithKline, and the Office of Research and Development, Biomedical Laboratory Research Program, US Department of Veterans Affairs Administration. For the ADGC, biological samples and associated phenotypic data used in primary data analyses were stored at principal investigators' institutions and at the National Cell Repository for Alzheimer's Disease (NCRAD) at Indiana University, funded by the NIA. Associated phenotypic data used in secondary data analyses were stored at the National Alzheimer's Coordinating Center and at the NIA Alzheimer's Disease Data Storage Site at the University of Pennsylvania, funded by the NIA. Contributors to the genetic analysis data included principal investigators on projects ndividually funded by the NIA, other NIH institutes, or private entities.

**Acknowledgments for other GWAS and phenotype data**

*NACC*

The NACC database is funded by NIA/NIH Grant U01 AG016976. NACC data are contributed by the NIA-funded ADCs: P30 AG019610 (PI Eric Reiman, MD), P30 AG013846 (PI Neil Kowall, MD), P30 AG062428-01 (PI James Leverenz, MD) P50 AG008702 (PI Scott Small, MD), P50 AG025688 (PI Allan Levey, MD, PhD), P50 AG047266 (PI Todd Golde, MD, PhD), P30 AG010133 (PI Andrew Saykin, PsyD), P50 AG005146 (PI Marilyn Albert, PhD), P30 AG062421-01 (PI Bradley Hyman, MD, PhD), P30 AG062422-01 (PI Ronald Petersen, MD, PhD), P50 AG005138 (PI Mary Sano, PhD), P30 AG008051 (PI Thomas Wisniewski, MD), P30 AG013854 (PI Robert Vassar, PhD), P30 AG008017 (PI Jeffrey Kaye, MD), P30 AG010161 (PI David Bennett, MD), P50 AG047366 (PI Victor Henderson, MD, MS), P30 AG010129 (PI Charles DeCarli, MD), P50 AG016573 (PI Frank LaFerla, PhD), P30 AG062429-01(PI James Brewer, MD, PhD), P50 AG023501 (PI Bruce Miller, MD), P30 AG035982 (PI Russell Swerdlow, MD), P30 AG028383 (PI Linda Van Eldik, PhD), P30 AG053760 (PI Henry Paulson, MD, PhD), P30 AG010124 (PI John Trojanowski, MD, PhD), P50 AG005133 (PI Oscar Lopez, MD), P50 AG005142 (PI Helena Chui, MD), P30 AG012300 (PI Roger Rosenberg, MD), P30 AG049638 (PI Suzanne Craft, PhD), P50 AG005136 (PI Thomas Grabowski, MD), P30 AG062715-01 (PI Sanjay Asthana, MD, FRCP), P50 AG005681 (PI John Morris, MD), P50 AG047270 (PI Stephen Strittmatter, MD, PhD).

*MARS & LATC*

We thank all Minority Aging Research Study and Latino Core participants and the Rush Alzheimer's Disease Center staff. This database was funded by the NIH/NIA grants R01AG22018 (MARS) and P30AG 072975 (ADC).

*GenADA*

The genotypic and associated phenotypic data used in the study "Multi-Site Collaborative Study for Genotype-Phenotype Associations in Alzheimer's Disease (GenADA)" were provided by the GlaxoSmithKline, R&D Limited.

*ROSMAP*

ROSMAP study data were provided by the Rush Alzheimer's Disease Center, Rush University Medical Center, Chicago. Data collection was supported through funding by NIA grants P30AG10161, R01AG15819, R01AG17917, R01AG30146, R01AG36836, U01AG32984, U01AG46152, the Illinois Department of Public Health, and the Translational Genomics Research Institute.

*AddNeuroMed*

The AddNeuroMed data are from a public-private partnership supported by EFPIA companies and SMEs as part of InnoMed (Innovative Medicines in Europe), an Integrated Project funded by the European Union of the Sixth Framework program priority FP6-2004-LIFESCIHEALTH-5. Clinical leads responsible for data collection are Iwona Kłoszewska (Lodz), Simon Lovestone (London), Patrizia Mecocci (Perugia), Hilkka Soininen (Kuopio), Magda Tsolaki (Thessaloniki), and Bruno Vellas (Toulouse), imaging leads are Andy Simmons (London), Lars-Olad Wahlund (Stockholm) and Christian Spenger (Zurich) and bioinformatics leads are Richard Dobson (London) and Stephen Newhouse (London).

*ADNI*

Data collection and sharing for this project was funded by the Alzheimer's Disease Neuroimaging Initiative (ADNI) (National Institutes of Health Grant U01 AG024904) and DOD ADNI (Department of Defense award number W81XWH-12-2-0012). ADNI is funded by the National Institute on Aging, the National Institute of Biomedical Imaging and Bioengineering and through generous contributions from the following: AbbVie. Alzheimer's Association; Alzheimer's Drug Discovery Foundation; Araclon Biotech; BioClinica. Inc.; Biogen; Bristol-Myers Squibb Company; CereSpir. Inc.; Cogstate; Eisai Inc.; Elan Pharmaceuticals. Inc.; Eli Lilly and Company; EuroImmun; F. Hoffmann-La Roche Ltd and its affiliated company Genentech. Inc.; Fujirebio; GE HealtControlsare; IXICO Ltd.; Janssen Alzheimer Immunotherapy Research & Development. LLC.; Johnson & Johnson Pharmaceutical Research & Development LLC.; Lumosity; Lundbeck; Merck & Co. Inc.; Meso Scale Diagnostics. LLC.; NeuroRx Research; Neurotrack Technologies; Novartis Pharmaceuticals Corporation; Pfizer Inc.; Piramal Imaging; Servier; Takeda Pharmaceutical Company; and Transition Therapeutics. The Canadian Institutes of Health Research is providing funds to support ADNI clinical sites in Canada. Private sector contributions are facilitated by the Foundation for the National Institutes of Health. The grantee organization is the Northern California Institute for Research and Education, and the study is coordinated by the Alzheimer's Therapeutic Research Institute at the University of Southern California. ADNI data are disseminated by the Laboratory for Neuro Imaging at the University of Southern California.

*NCRAD*

Biological samples used in this study were stored at study investigators' institutions and at the National Cell Repository for Alzheimer's Disease (NCRAD) at Indiana University, which receives government support under a cooperative agreement grant (U24 AG21886) awarded by the National Institute on Aging (NIA). We thank contributors who collected samples used in this study, as well as patients and their families, whose help and participation made this work possible.

*UK Biobank*

UK Biobank data were analyzed under Application Number 45420.

*FinnGen Study*

We want to acknowledge the participants and investigators of the FinnGen study. The FinnGen project is funded by two grants from Business Finland (HUS 4685/31/2016 and UH 4386/31/2016) and the following industry partners: AbbVie Inc., Alnylam Pharmaceuticals, Inc., AstraZeneca UK Ltd, Bayer AG, Biogen MA Inc., Boehringer Ingelheim International GmbH, Bristol Myers Squibb Inc. (and Celgene Corporation & Celgene International II Sàrl), Genentech Inc., GlaxoSmithKline Intellectual Property Development Ltd., Johnson&Johnson Innovative Medicine Inc., Maze Therapeutics Inc., Merck Sharp & Dohme LCC, Novartis AG, Pfizer Inc. and Sanofi US Services Inc. Following biobanks are acknowledged for delivering biobank samples to FinnGen: Auria Biobank (www.auria.fi/biopankki), THL Biobank (www.thl.fi/biobank), Helsinki Biobank (www.helsinginbiopankki.fi), Biobank Borealis of Northern Finland (https://www.ppshp.fi/Tutkimus-ja-opetus/Biopankki/Pages/Biobank-Borealis-briefly-in-English.aspx), Finnish Clinical Biobank Tampere (www.tays.fi/en-US/Research_and_development/Finnish_Clinical_Biobank_Tampere), Biobank of Eastern Finland (www.ita-suomenbiopankki.fi/en), Central Finland Biobank (www.ksshp.fi/fi-FI/Potilaalle/Biopankki), Finnish Red Cross Blood Service Biobank (www.veripalvelu.fi/verenluovutus/biopankkitoiminta), Terveystalo Biobank (www.terveystalo.com/fi/Yritystietoa/Terveystalo-Biopankki/Biopankki/) and Arctic Biobank (https://www.oulu.fi/en/university/faculties-and-units/faculty-medicine/northern-finland-birth-cohorts-and-arctic-biobank). All Finnish Biobanks are members of BBMRI.fi infrastructure (https://www.bbmri-eric.eu/national-nodes/finland/). Finnish Biobank Cooperative -FINBB (https://finbb.fi/) is the coordinator of BBMRI-ERIC operations in Finland. The Finnish biobank data can be accessed through the Fingenious® services (https://site.fingenious.fi/en/) managed by FINBB.

*FinnGen ethics statement*

Study subjects in FinnGen provided informed consent for biobank research, based on the Finnish Biobank Act. Alternatively, separate research cohorts, collected prior the Finnish Biobank Act came into effect (in September 2013) and start of FinnGen (August 2017), were collected based on study-specific consents and later transferred to the Finnish biobanks after approval by Fimea (Finnish Medicines Agency), the National Supervisory Authority for Welfare and Health. Recruitment protocols followed the biobank protocols approved by Fimea. The Coordinating Ethics Committee of the Hospital District of Helsinki and Uusimaa (HUS) statement number for the FinnGen study is Nr HUS/990/2017.

The FinnGen study is approved by Finnish Institute for Health and Welfare (permit numbers: THL/2031/6.02.00/2017, THL/1101/5.05.00/2017, THL/341/6.02.00/2018, THL/2222/6.02.00/2018, THL/283/6.02.00/2019, THL/1721/5.05.00/2019 and THL/1524/5.05.00/2020), Digital and population data service agency (permit numbers: VRK43431/2017-3, VRK/6909/2018-3, VRK/4415/2019-3), the Social Insurance Institution (permit numbers: KELA 58/522/2017, KELA 131/522/2018, KELA 70/522/2019, KELA 98/522/2019, KELA 134/522/2019, KELA 138/522/2019, KELA 2/522/2020, KELA 16/522/2020), Findata permit numbers THL/2364/14.02/2020, THL/4055/14.06.00/2020, THL/3433/14.06.00/2020, THL/4432/14.06/2020, THL/5189/14.06/2020, THL/5894/14.06.00/2020, THL/6619/14.06.00/2020, THL/209/14.06.00/2021, THL/688/14.06.00/2021, THL/1284/14.06.00/2021, THL/1965/14.06.00/2021, THL/5546/14.02.00/2020, THL/2658/14.06.00/2021, THL/4235/14.06.00/2021, Statistics Finland (permit numbers: TK-53-1041-17 and TK/143/07.03.00/2020 (earlier TK-53-90-20) TK/1735/07.03.00/2021, TK/3112/07.03.00/2021) and Finnish Registry for Kidney Diseases permission/extract from the meeting minutes on 4$^{th}$ July 2019.

The Biobank Access Decisions for FinnGen samples and data utilized in FinnGen Data Freeze 12 include: THL Biobank BB2017_55, BB2017_111, BB2018_19, BB_2018_34, BB_2018_67, BB2018_71, BB2019_7, BB2019_8, BB2019_26, BB2020_1, BB2021_65, Finnish Red Cross Blood Service Biobank 7.12.2017, Helsinki Biobank HUS/359/2017, HUS/248/2020, HUS/430/2021 §28, §29, HUS/150/2022 §12, §13, §14, §15, §16, §17, §18, §23, §58, §59, HUS/128/2023 §18, Auria Biobank AB17-5154 and amendment #1 (August 17 2020) and amendments BB_2021-0140, BB_2021-0156 (August 26 2021, Feb 2 2022), BB_2021-0169, BB_2021-0179, BB_2021-0161, AB20-5926 and amendment #1 (April 23 2020) and it´s modifications (Sep 22 2021), BB_2022-0262, BB_2022-0256, Biobank Borealis of Northern Finland_2017_1013, 2021_5010, 2021_5010 Amendment, 2021_5018, 2021_5018 Amendment, 2021_5015, 2021_5015 Amendment, 2021_5015 Amendment_2, 2021_5023, 2021_5023 Amendment, 2021_5023 Amendment_2, 2021_5017, 2021_5017 Amendment, 2022_6001, 2022_6001 Amendment, 2022_6006 Amendment, 2022_6006 Amendment, 2022_6006 Amendment_2, BB22-0067, 2022_0262, 2022_0262 Amendment, Biobank of Eastern Finland 1186/2018 and amendment 22§/2020, 53§/2021, 13§/2022, 14§/2022, 15§/2022, 27§/2022, 28§/2022, 29§/2022, 33§/2022, 35§/2022, 36§/2022, 37§/2022, 39§/2022, 7§/2023, 32§/2023, 33§/2023, 34§/2023, 35§/2023, 36§/2023, 37§/2023, 38§/2023,

39§/2023, 40§/2023, 41§/2023, Finnish Clinical Biobank Tampere MH0004 and amendments (21.02.2020 & 06.10.2020), BB2021-0140 8§/2021, 9§/2021, §9/2022, §10/2022, §12/2022, 13§/2022, §20/2022, §21/2022, §22/2022, §23/2022, 28§/2022, 29§/2022, 30§/2022, 31§/2022, 32§/2022, 38§/2022, 40§/2022, 42§/2022, 1§/2023, Central Finland Biobank 1-2017, BB_2021-0161, BB_2021-0169, BB_2021-0179, BB_2021-0170, BB_2022-0256, BB_2022-0262, BB22-0067, Decision allowing to continue data processing until 31st Aug 2024 for projects: BB_2021-0179, BB22-0067,BB_2022-0262, BB_2021-0170, BB_2021-0164, BB_2021-0161, and BB_2021-0169, and Terveystalo Biobank STB 2018001 and amendment 25th Aug 2020, Finnish Hematological Registry and Clinical Biobank decision 18th June 2021, Arctic biobank P0844: ARC_2021_1001.
